# Heterogeneity, Longitudinal Decline, and Metabolic Risk in MRI-Based Quantification of 20 Individual Hip and Thigh Muscles

**DOI:** 10.64898/2026.02.25.26347009

**Authors:** Brandon Whitcher, Hamzah Raza, Nicolas Basty, Marjola Thanaj, Camilo Bell-Bradford, Marili Niglas, Jimmy D. Bell, E. Louise Thomas, Dimitri Amiras

## Abstract

Quantifying muscle health at scale has been limited by the difficulty of segmenting individual muscles on MRI. We developed an automated 3D deep-learning framework that segments 20 bilateral hip and thigh muscles from Dixon MRI, enabling muscle level quantification of volume and relative fat fraction (rFF). Applied to 10,840 baseline and 2,766 longitudinal UK Biobank scans, this framework supports population-scale phenotyping across demographic, metabolic and treatment exposures. Segmentation accuracy was robust, and increased with muscle size. Men had greater muscle volumes, whereas women showed consistently higher rFF. Fat infiltration was highest in postural and pelvic-stabilising muscles and lowest in the quadriceps, revealing pronounced anatomical heterogeneity. Over two years, most muscles showed small but consistent volume declines, with losses more uniform in men and more heterogeneous in women; rFF increased more prominently in women, suggesting early compositional deterioration. In type 2 diabetes, men showed widespread volume loss and elevated rFF, whereas women showed minimal volume loss and heterogeneous fat changes, revealing sex-specific disease signatures. Automated muscle-specific MRI phenotyping resolves structural and compositional changes obscured by compartment-level measures and provides a scalable platform for population-level studies of musculoskeletal ageing, metabolic disease, and therapeutic response.

## Introduction

The importance of muscle mass, strength and quality are increasingly recognised as having a central role in health, ageing and metabolic disease. Lower skeletal muscle mass reduces whole-body capacity for glucose uptake^1^, while greater intramuscular fat infiltration disrupts insulin signalling^2^; making these central determinants of type-2 diabetes (T2D) risk and progression^3,4^ and predictors of incident T2D in large population cohorts^5^. These metabolic consequences sit within a broader landscape of musculoskeletal ageing. Sarcopenia, the progressive and generalised loss of skeletal muscle mass and strength, is strongly associated with physical disability, reduced quality of life, and increased mortality^6,7^. Yet there is still little clear understanding regarding why some muscles deteriorate earlier or more severely than others during ageing or disease^8^. Proposed explanations include the selective loss and atrophy of particular fibre types and age-related remodelling of fibre phenotype^9,10^, however, confirming these mechanisms relies on muscle biopsy, challenging in the deeper muscle groups, making them impossible to investigate in large cohorts.

Skeletal asymmetry is well known, with directional asymmetry observed in both the upper and lower limbs, most commonly due to behavioural changes. Whilst the right upper limb is often noted to be larger in right-handed individuals than the left lower limb, this can be reversed^11^. Directional asymmetry is also noted with neurodegenerative conditions such as Parkinson’s disease^12^ and there is some evidence to suggest asymmetry could increase risk of injury in sport^13^.

Selective vulnerability is even more apparent in rare neuromyopathic conditions, where complex genetic interactions along the neuromuscular pathway produce strikingly patterned muscle involvement^14,15^. Certain muscles undergo rapid atrophy or fat infiltration while neighbouring muscles remain relatively preserved, revealing biological distinctions that conventional imaging approaches often fail to capture. In addition to the distinct fibre-type composition of individual muscles, habitual loading patterns differ markedly with gait, posture, and physical activity, shape muscle-specific susceptibility to disuse, injury, and metabolic decline^16,17^. Clinically, the functional consequences of muscle loss are also muscle-specific: atrophy of hip abductors affects balance and fall risk, quadriceps loss impairs mobility and stair climbing, and hamstring deterioration alters gait stability^18,19^. Sex differences in muscle distribution, fibre composition, and fat infiltration further reinforce the need for granular, muscle-level analysis^20^.

Heterogeneity of muscle involvement in disease highlights a fundamental gap: imaging-based assessments of muscle health rely on generalised area or volumetric measures derived from single slices, multiple slices/slabs or total thigh/lower leg volumes^21,22^. Alternative approaches include subdividing these volumes into simple anterior/posterior compartment divisions^23^ or functional groups^24,25^. This has traditionally been related to the complexity and time required to manually annotate individual thigh muscles on magnetic resonance imaging (MRI) or computed tomography (CT) images. Fuchs *et al*.^26^ estimated it took approximately 1,600 hours to manually segment thigh and calf images in only 20 participants. Advances in image segmentation and the application of deep learning methods have transformed image-based measurement of body composition^27^, enabling measurement of total and regional muscle volumes at scale. To address these limitations, we and others have developed automated image-analysis methods capable of quantifying the volume^28,29^ or both volume and fat content of individual leg muscles^30,31^. However the number of muscles that the different published models are able to segment varies substantially.

Leg muscle segmentation in MRI has evolved from semi-automated methods^32^, iterative thresholding^33^, and non-linear registration propagation^34^. Methodology has shifted towards two-dimensional (2D) deep learning approaches^28,35^, statistical shape models^36^, or hybrid ensembles that have been shown to outperform standard 3D UNet benchmarks^25^. Approaches have also utilised 3D-to-2D parametric mapping^37^, as well as 3D and multi-stage pipelines ranging from multi-atlas fusion for major muscle groups^38^ to 3D attention-based models, two-stage networks targeting a comprehensive set of muscles across the thigh and lower leg^31,39^, and transformer-based architectures for 3D muscle segmentation^40^.

In this paper, we present a deep-learning method to measure the volume and fat content of 20 hip and thigh muscles from MRI scans collected in the UK Biobank. Using these tools, we examined sex-specific differences in muscle morphology, mapped longitudinal patterns of muscle loss, explored how distinct muscles are affected by impaired metabolic regulation and assessed regional asymmetry in muscle volumes. We also identified participants who reported using GLP-1RA, given emerging evidence that these agents can induce substantial weight loss, including reductions in lean mass, raising important questions about their effects on muscle quality, metabolic function, and vulnerability to sarcopenic decline^41^. By focusing on the variation that is neglected in compartment-level analyses, this approach opens new opportunities to understand the pathways linking musculoskeletal decline, metabolic disease, and neuromuscular vulnerability, and to identify more targeted biomarkers and intervention points.

## Methods

### Data

The UK Biobank recruited over 500,000 participants from across the UK, aged 40 to 85, between 2006 and 2014^42^, with a subset of 100,000 participants selected to undergo MRI of the heart, brain, and abdomen^43^. The UK Biobank study was approved by the North West Multi-centre Research Ethics Committee (REC reference: 11/NW/0382) with informed consent from all participants. Researchers may apply to use the UK Biobank data resource by submitting a health-related research proposal in the public interest. All methods were performed in accordance with the relevant guidelines and regulations as presented by the appropriate authorities.

### Phenotype definitions

Anthropometric and lifestyle measurements, including age, weight, height, waist circumference, and hip circumference, systolic and diastolic blood pressure, hand grip strength (HGS) in the dominant hand, metabolic equivalent task (MET), alcohol intake and smoking status, were acquired at the UK Biobank imaging visit. Body mass index (BMI) and waist-to-hip ratio were calculated from them. Sedentary time was quantified as the total daily time (hours/day) spent watching television, using a computer, and driving, derived from self-reported touchscreen questionnaire responses at the imaging visits, with values > 16 hours/day excluded^44^. Ethnicity was defined based on self-reported genetic background at the initial assessment. The Townsend deprivation index was calculated immediately prior to participants attending their initial assessment.

Type-2 diabetes (T2D) was defined based on the International Classification of Diseases 10th edition (ICD-10) for T2D using E11, self-reported codes 1220 or 1223, respectively^45^. GLP-1RA medication codes were identified from primary care prescription records using drug-name matching for GLP-1RA substances, including exenatide, liraglutide, lixisenatide, albiglutide, dulaglutide, semaglutide, beinaglutide, and tirzepatide^46,47^. Prescriptions issued prior to the imaging visit were retained and linked to participant identifiers. Duration of exposure was calculated as the time between the first recorded prescription and the imaging visit date. Participants were classified as GLP-1RA users if the mean exposure duration exceeded six months.

### Image acquisition and processing

We utilised the 3D two-point Dixon sequences, which gives coverage from neck to knees, from 11,503 participants in the UK Biobank. Comprehensive details on the MRI abdominal protocol have been reported previously^43^. A total of 663 participants were excluded from our final analysis due to various issues that directly affected the quality of the leg-muscle segmentations. These included an implant in at least one hip or knee (*n* = 42), the head in the acquisition (resulting in the legs only being partially covered, *n* = 2), no leg images (*n* = 4), extreme fat infiltration (*n* = 14), and poor image acquisition or reconstruction (*n* = 22). While we acknowledge that the segmentation model will be used to provide fat quantification, the extreme levels of fat infiltration observed in the 14 participants prevented individual muscle groups from being identified (see Figure 2B for an example). Because the Dixon acquisition was fixed with respect to its superior-inferior coverage (at 1.1 m), complete coverage of the upper legs was not possible for taller participants, resulting in underestimation of some muscle groups. Therefore, participants with a height > 1.85 m were excluded (*n* = 506). Finally, to ensure all participants had both muscle volume and fat fraction measurements, individuals without measurable fat fraction were also excluded (*n* = 73). Thus, a total of 10,840 baseline scans from unique participants were utilised, with re-imaging scans available for 2,766 of these participants.

All data were pre-processed using established automated pipelines^27,48^. Specifically, the Dixon series were resampled to a fixed dimension and resolution to facilitate merging the six series into a single neck-to-knee volume (size = [224, 174, 370], resolution = 2.232 × 2.232 × 3.0 mm^3^). We identified a fixed set of slices that form an overlap (in the inferior-superior direction) between adjacent series and applied a nonlinear function to blend the signal intensities, where slices in the interior of the volume were more heavily weighted and slices near the boundary were suppressed. For leg segmentation, 20 bilateral muscle groups (Figure 1) were annotated in 131 participants using the opposed-phase channel from the Dixon MRI. Of these, 28 datasets were fully annotated manually. A further seven datasets were manually refined from a deep-learning model trained on these initial datasets. Fifty-nine datasets were accepted without manual editing following expert visual inspection, based on a model trained on the first 35 manually annotated cases. This ground-truth set (*n* = 94) was used to train a final model, from which an additional 32 datasets were accepted without manual editing, and five underweight cases (BMI < 18.5 kg/m^2^) were manually refined, yielding a total of 131 ground-truth datasets. Some muscles were too small to be differentiated given the resolution of the Dixon MRI, these were combined into “pelvic floor” and “short external rotator” muscle groups.

**Figure 1.**
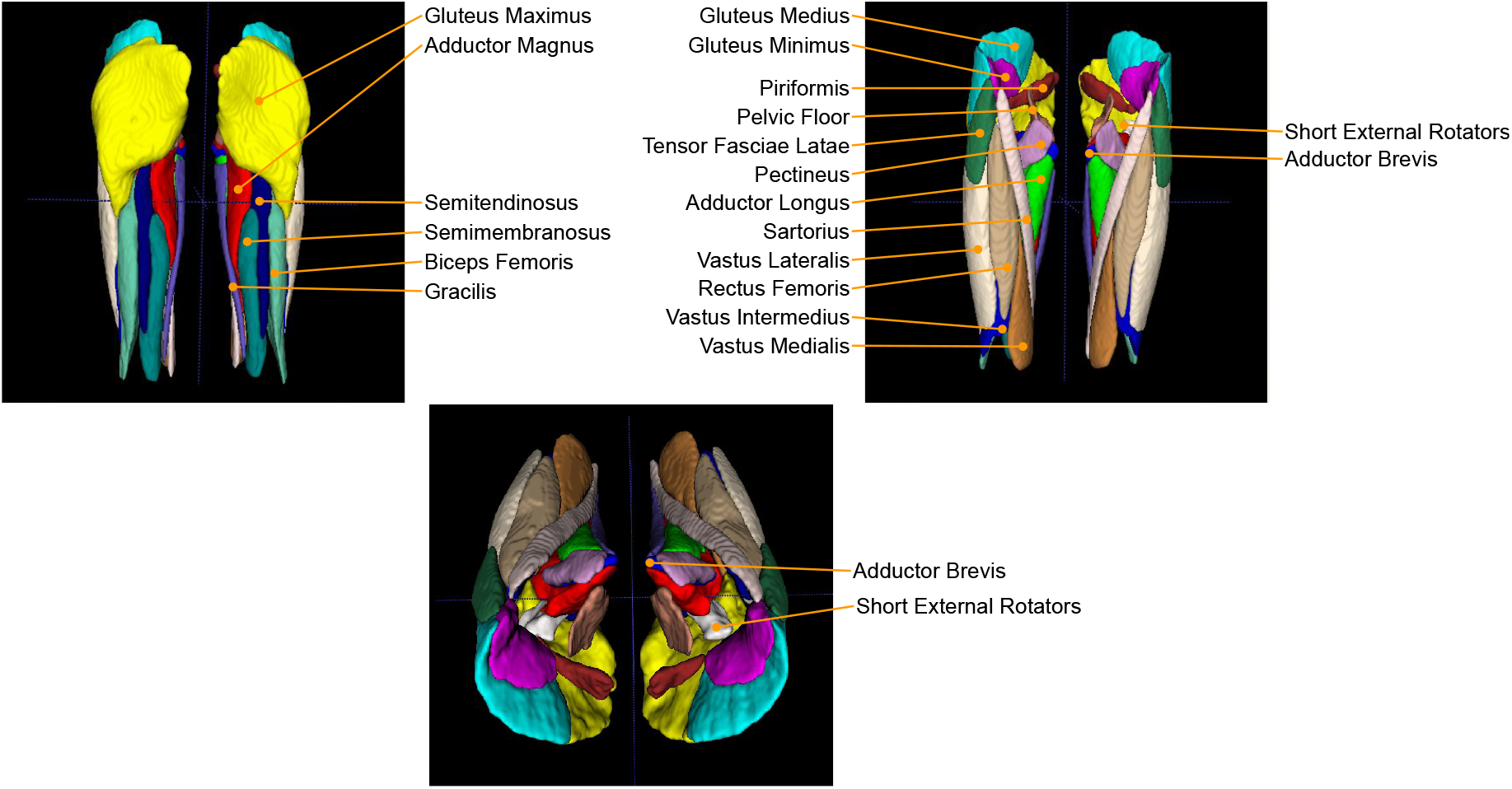
Twenty bilateral muscle groups covering the hips and thighs. The short external rotators are composed of the following muscle groups: gemellus superior, obturator internus, gemellus inferior, obturator externus, and quadratus femoris.

The ground-truth data were split 95:5 for training and testing the deep learning segmentation model, respectively. The test datasets contained a mixture of 100% manual annotations, model-generated output, and manually-edited model output. The relatively small number of test datasets was chosen to balance the fact that only a modest number of manual annotations were available. The Swin-UNETR model architecture was selected based on its performance in segmenting complex collections of multiple labels in medical imaging data^49^. It was implemented using MONAI^50^ with weights from self-supervised pre-training datasets^51^. Data augmentation was performed and a total of 45,000 steps executed, with the final model selected using the Dice score on out-of-sample test datasets.

Taking advantage of the information contained in Dixon MRI, we computed the rFF in the 20 bilateral muscle groups. Relative fat fraction is defined to be the fat signal divided by the sum of the fat and water signals in a single voxel. This produces a measure of muscle quality through estimating the median rFF over all voxels in the segmented muscles.

### Statistical analysis

All summary statistics, hypothesis tests and figures were performed using the R software environment for statistical computing and graphics^52^. Descriptive statistics for continuous variables are expressed as mean and standard deviation. The Wilcoxon rank-sum test was used to compare means between groups, and the Wilcoxon signed-rank test with paired observations. The threshold for statistical significance of *p*-values was adjusted for the number of formal hypothesis tests performed in Supplementary Tables S3, S4, S6, S7, S9 and S10. The Bonferroni-corrected threshold was 0.05*/*480 = 0.00010417.

To compare muscle volume and rFF image-derived phenotypes (IDPs) between participants with and without T2D, we identified the participants with T2D who were diagnosed before the baseline imaging visit. We then identified a control cohort without any reported conditions and designed a case-control study for each disease population, ultimately achieving a case-control cohort with T2D. The sex-stratified control cohort was selected by matching each case with one control by age (within ±2 years) and BMI (within ±1.5 kg/m^2^).

To examine muscle volume and rFF IDPs between participants prescribed with GLP-1RA, a total of 38 participants prescribed with GLP-1RA for at least six months before the baseline imaging visit (mean duration 5.9 ± 2.9 years, range 0.7 − 12 years) were matched by sex, age (±1 years), BMI (±1.5 kg/m^2^), Townsend deprivation index (±1), T2D, hypertension, cardiovascular disease, muscle disorders, sedentary lifestyle (> 10.6 hours/day), and cholesterol medication with non-GLP-1RA controls.

## Results

### Model performance and quality control

The average Dice score over all test datasets and muscle groups was 0.871. The average Dice scores for individual muscle groups varied between 0.742 for the adductor brevis and 0.940 for the gluteus maximus (Supplementary Figure S1). Some participants were excluded from the quantitative segmentation analysis because the automated pipeline could not reliably delineate individual muscles. However, we retained these cases in the qualitative results as the underlying reasons for segmentation failure, typically marked pathology or substantial artefact that distorts normal muscle morphology, are themselves clinically meaningful. In a real-world workflow, such cases would be flagged for targeted visual review, since they often represent important abnormalities that warrant further clinical attention.

### Participants

A total of 10,840 baseline scans from participants in the UK Biobank were analyzed. Of these, 1,716 participants had a diagnosis of T2D (with case controls), while a further 38 participants were prescribed GLP-1RA for more than six months. The remaining subjects were a representative population of the general UK Biobank imaging cohort. Table 1 summarises the participant demographics of the study population, of whom 86% had white ethnicity, 48% were women, and the overall cohort had an average age of 65 ± 7 years. Physical and anthropometric variables differed between sexes with men having a higher waist circumference (men: 96 ±12 cm; women: 85 ±14 cm), visceral adipose tissue (VAT) volume (men: 5.87 ± 2.56 l; women: 3.25 ± 1.83 l), and HGS (men: 38 ± 9 kg; women: 24 ± 6 kg). T2D was more prevalent among men (24% versus 14% for women).

**Table 1.** Demographics for participants in the overall cohort, separated by sex. Values for continuous variables are reported as mean and standard deviation, values for discrete variables are reported as percentages. ASAT: abdominal subcutaneous adipose tissue; BMI: body mass index; GLP-1 RA: glucagon-like peptide-1 receptor agonist; MET: metabolic equivalent of task; T2D: type-2 diabetes; VAT: visceral adipose tissue.

| Characteristic | Women<br>*n* = 5,199<sup>†</sup> | Men<br>*n* = 5,641<sup>†</sup> | Overall<br>*n* = 10,840<sup>†</sup> |
| --- | --- | --- | --- |
| Age (years) | 64 (7) | 66 (7) | 65 (7) |
| White Ethnicity | 4,441 (85%) | 4,933 (87%) | 9,374 (86%) |
| Weight (kg) | 71 (15) | 85 (14) | 78 (16) |
| Height (m) | 1.62 (0.06) | 1.75 (0.06) | 1.69 (0.09) |
| BMI (kg/m<sup>2</sup>) | 26.8 (5.6) | 27.8 (4.4) | 27.3 (5.0) |
| Waist Circumference (cm) | 85 (14) | 96 (12) | 91 (14) |
| Hip Circumference (cm) | 102 (11) | 102 (8) | 102 (10) |
| Waist-to-Hip Ratio | 0.83 (0.07) | 0.95 (0.07) | 0.89 (0.09) |
| Dominant Hand Grip Strength (kg) | 24 (6) | 38 (9) | 31 (10) |
| Systolic Blood Pressure (mmHg) | 137 (19) | 143 (17) | 140 (18) |
| Diastolic Blood Pressure (mmHg) | 77 (10) | 80 (10) | 79 (10) |
| Total MET (hours/week) | 49 (42) | 48 (43) | 48 (42) |
| Sedentary Time (hours) | 5.07 (2.16) | 5.78 (2.34) | 5.44 (2.28) |
| GLP-1RA Intake | 20 (0.4%) | 48 (0.9%) | 68 (0.6%) |
| T2D | 722 (14%) | 1,340 (24%) | 2,062 (19%) |
| ASAT Volume (l) | 10.7 (5.1) | 7.8 (3.8) | 9.2 (4.7) |
| VAT Volume (l) | 3.25 (1.83) | 5.87 (2.56) | 4.61 (2.59) |

<sup>†</sup>Mean (SD); *n* (%)

### Muscle asymmetry in the overall cohort

Our segmentation model enabled the investigation of left-right symmetry in muscle volumes. Supplementary Figures S2 and S3 show the relationships between left and right muscles for men and women, respectively, and a summary of the linear regression models is provided in in Supplementary Table S11. Across muscle groups, left and right volumes were strongly correlated, with most data points closely aligned along the line of identity. However, for several muscles, the association was weaker. This pattern was observed in both men and women for a variety of smaller muscle groups such as adductor brevis, gluteus minimus, pelvic floor, piriformis and short external rotators.

As well as providing useful cohort information, this method is applicable at the individual level, enabling identification of localized leg muscle asymmetries. Figure 2 provides three examples of asymmetries in UK Biobank participants: one with severe fatty change in the left thigh and upper calf, one shows similar replacement of the normal muscle of the left thigh with fat and some of the left upper calf, and one with an extreme left-right asymmetry involving only muscle groups in the left hip.

**Figure 2.**
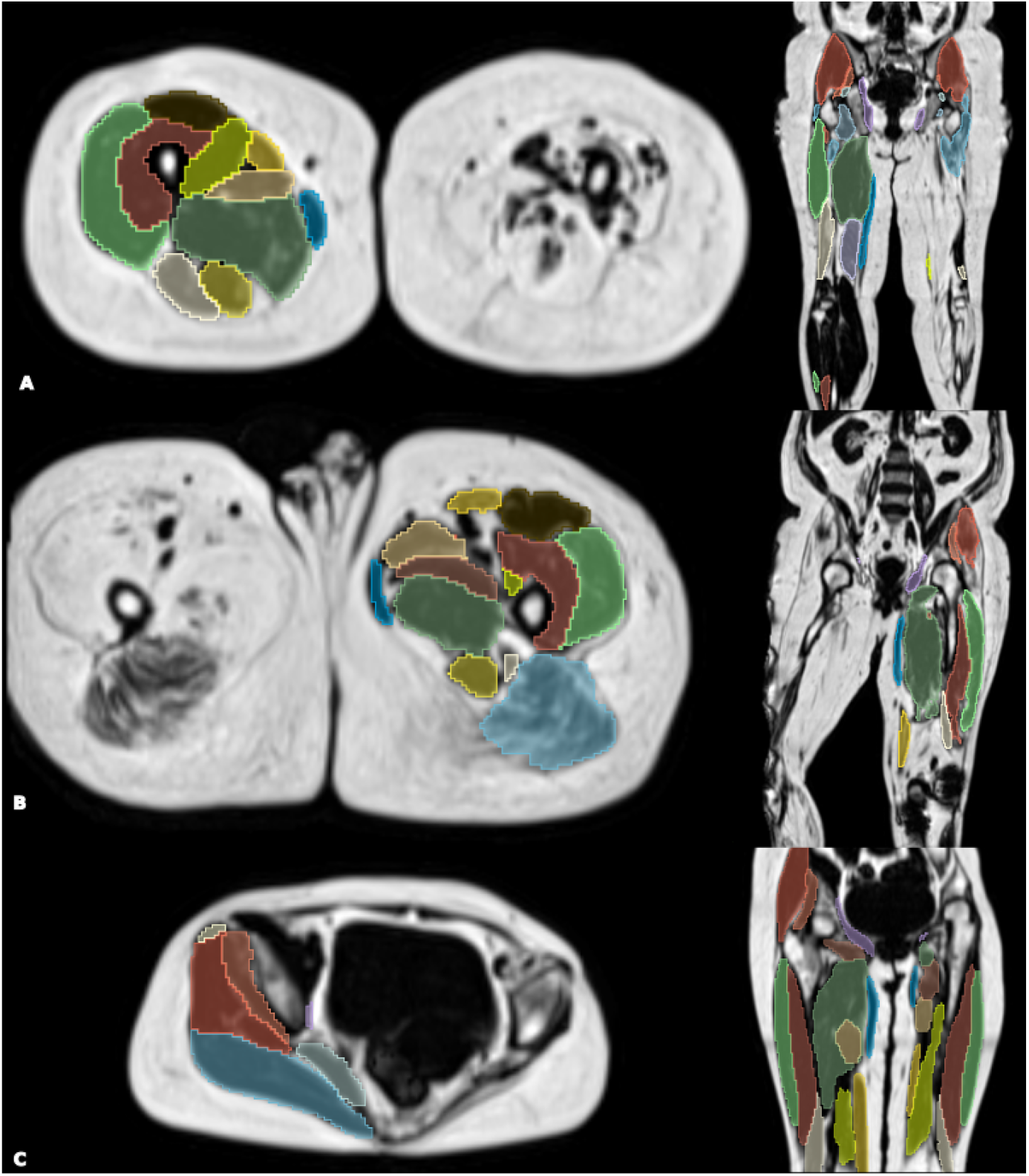
Axial and coronal Dixon MRI (fat channel) of three participants with marked muscle asymmetry. Participant A shows severe fatty change in the left thigh and upper calf. Participant B shows similar replacement of normal muscle in the left thigh and upper calf with fat. Participant C demonstrates severe atrophy of the left gluteus muscles and iliopsoas with associated left hip dysplasia with fatty infiltration and marked volume loss of the left gluteus maximus. All cases are suggestive of a neurogenic disease with previous poliomyelitis infection. Reproduced by kind permission of UK Biobank ©.

All three participants suggest a neurogenic cause, the most likely of which is poliomyelitis infection. Identifying cases like this may provide useful insights into distinguishing between pathological atrophy and normal variations.

### Sex differences in muscle volume and fat fraction

The volumes of the 20 bilateral muscle groups for all participants (separated by sex) are summarised in Table 2. Substantial variation was noted across muscle groups, with largest absolute volumes observed for the gluteus maximus (1, 680 ± 394 ml), the adductor magnus (1, 133 ± 293 ml) and the vastus lateralis (927 ± 238 ml). All muscle volumes were consistently higher in men. Smaller absolute volume differences between sexes were seen in muscles such as the piriformis (91 ± 16 ml; 86 ± 17 ml), the pectineus (104 ± 16 ml; 97 ± 18 ml), and the short external rotators (men: 103± 21 ml; women: 87 ±19 ml).

**Table 2.** Total volume, in milliliters, of the twenty bilateral muscle groups in the hips and thighs, separated by sex. Values are reported as mean and standard deviation.

| Image-Derived Phenotype | Women<br>n = 5,199<sup>†</sup> | Men<br>n = 5,641<sup>†</sup> | Overall<br>n = 10,840<sup>†</sup> |
| --- | --- | --- | --- |
| Adductor Brevis (ml) | 106 (22) | 165 (28) | 137 (39) |
| Adductor Longus (ml) | 244 (54) | 321 (56) | 284 (68) |
| Adductor Magnus (ml) | 903 (166) | 1,346 (213) | 1,133 (293) |
| Biceps Femoris (ml) | 462 (84) | 655 (106) | 562 (136) |
| Gluteus Maximus (ml) | 1,427 (276) | 1,914 (337) | 1,680 (394) |
| Gluteus Medius (ml) | 573 (86) | 750 (103) | 666 (130) |
| Gluteus Minimus (ml) | 152 (30) | 199 (34) | 176 (39) |
| Gracilis (ml) | 156 (36) | 208 (41) | 183 (47) |
| Pectineus (ml) | 97 (18) | 104 (16) | 100 (17) |
| Pelvic Floor (ml) | 117 (16) | 131 (16) | 124 (17) |
| Piriformis (ml) | 86 (17) | 91 (16) | 88 (17) |
| Rectus Femoris (ml) | 331 (57) | 469 (83) | 403 (99) |
| Sartorius (ml) | 247 (51) | 318 (53) | 284 (63) |
| Semimembranosus (ml) | 359 (70) | 503 (92) | 434 (109) |
| Semitendinosus (ml) | 239 (47) | 338 (63) | 290 (75) |
| Short External Rotators (ml) | 87 (19) | 103 (21) | 95 (21) |
| Tensor Fasciae (ml) | 123 (35) | 156 (36) | 140 (39) |
| Vastus Intermedius (ml) | 653 (116) | 966 (160) | 816 (210) |
| Vastus Lateralis (ml) | 745 (137) | 1,095 (181) | 927 (238) |
| Vastus Medialis (ml) | 566 (99) | 798 (129) | 687 (164) |

<sup>†</sup>Mean (SD)

To ensure these sex differences represented muscle variation and were not simply a reflection of men generally being taller than women, muscle volume divided by the participant’s height squared were calculated (Supplementary Table S1). Overall, sex-related differences were reduced following height normalisation, although the majority of muscle groups remained larger in men. Several smaller muscle groups showed comparable or lower height-indexed volumes in men, including the pectineus (men: 33.9 ± 5.0 ml/m^2^; women: 36.6 ± 6.6 ml/m^2^), the pelvic floor (men: 42.8 ± 4.6 ml/m^2^; women: 44.1 ± 5.0 ml/m^2^), and the piriformis (men: 29.7 ± 5.0 ml/m^2^; women: 32.4 ± 6.3 ml/m^2^).

In addition, we determined the rFF in all 20 bilateral muscle groups for all participants and separately by sex (Table 3). Among hip muscles, higher average rFF was consistently observed in the tensor fasciae latae (19.9 ± 10.5%) and the gluteus maximus (18.2 ± 9.5%), and the sartorius (15.3 ± 7.9%). In contrast, rFFs were lower (< 7%) in the adductor group and across the quadriceps, including the vastus intermedius, vastus medialis and vastus lateralis. Relative fat fraction was generally higher in women than in men. Pronounced differences were observed in the gracilis (men: 9.1 ± 4.1%; women: 18.9 ± 7.4%), the sartorius (men: 11.1 ± 5.4%; women: 19.9 ± 7.6%), and tensor fasciae latae (men: 15.8 ± 8.3%; women: 24.3 ± 10.8%).

**Table 3.** Median relative fat fraction (rFF) of the 20 bilateral muscle groups in the hips and thighs, in all participants and also separated by sex. Values are reported as mean and standard deviation.

| Image-Derived Phenotype | Women<br>n = 5,199<sup>l</sup> | Men<br>n = 5,641<sup>l</sup> | Overall<br>n = 10,840<sup>l</sup> |
| --- | --- | --- | --- |
| Adductor Brevis (%) | 8.4 (3.50) | 5.3 (1.62) | 6.8 (3.11) |
| Adductor Longus (%) | 6.2 (2.34) | 6.0 (2.71) | 6.1 (2.54) |
| Adductor Magnus (%) | 6.6 (2.96) | 5.8 (2.48) | 6.1 (2.75) |
| Biceps Femoris (%) | 10.6 (5.49) | 8.6 (4.44) | 9.6 (5.07) |
| Gluteus Maximus (%) | 19.9 (9.89) | 16.7 (8.87) | 18.2 (9.52) |
| Gluteus Medius (%) | 9.9 (4.94) | 9.0 (4.30) | 9.4 (4.64) |
| Gluteus Minimus (%) | 14.6 (8.34) | 9.1 (4.82) | 11.8 (7.28) |
| Gracilis (%) | 18.9 (7.38) | 9.1 (4.11) | 13.8 (7.67) |
| Pectineus (%) | 9.7 (3.26) | 6.3 (1.81) | 7.9 (3.12) |
| Pelvic Floor (%) | 17.3 (6.75) | 10.1 (4.13) | 13.5 (6.61) |
| Piriformis (%) | 18.5 (6.07) | 12.3 (4.62) | 15.2 (6.20) |
| Rectus Femoris (%) | 7.7 (3.41) | 5.6 (1.84) | 6.6 (2.90) |
| Sartorius (%) | 19.9 (7.55) | 11.1 (5.44) | 15.3 (7.87) |
| Semimembranosus (%) | 13.6 (7.83) | 11.5 (6.52) | 12.5 (7.26) |
| Semitendinosus (%) | 10.3 (4.99) | 8.0 (3.80) | 9.1 (4.56) |
| Short External Rotators (%) | 14.5 (7.06) | 8.0 (4.14) | 11.1 (6.60) |
| Tensor Fasciae (%) | 24.3 (10.75) | 15.8 (8.32) | 19.9 (10.47) |
| Vastus Intermedius (%) | 5.2 (2.53) | 4.8 (1.98) | 5.0 (2.27) |
| Vastus Lateralis (%) | 6.1 (3.22) | 5.0 (2.10) | 5.6 (2.75) |
| Vastus Medialis (%) | 5.8 (3.02) | 4.7 (2.11) | 5.2 (2.65) |

<sup>l</sup>Mean (SD)

### Longitudinal cohort

A total of 2,766 participants in the overall cohort had complete imaging data performed twice. The average duration between visits was 2.24 ± 0.12 years for men and 2.25 ± 0.12 years for women (range 2-2.96 years across all participants). The demographics of this cohort are described in Supplementary Table S2. The waist-to-hip ratio changed significantly in the longitudinal cohort, yet the magnitude of the difference was small. Blood pressure increased over time in both sexes, with systolic blood pressure rising from 135 to 139 mmHg in women and from 142 to 144 mmHg in men. Dominant HGS was reduced in both sexes (women: 24 to 22 kg; men: 39 to 36 kg). Physical activity levels were largely unchanged, while sedentary time increased in both women (4.89 to 5.03 hours/day) and men (5.47 to 5.63 hours/day). Visceral adipose tissue volume increased over time in both women (2.87 to 2.94 l) and men (5.21 to 5.27 l).

There were changes in individual muscle volumes at re-imaging; these are presented as absolute and relative percentage changes in Supplementary Table S3 separately by sex. Most muscle groups in men exhibited small but consistent reductions in volume. The largest decreases in volume were observed in major muscle groups such as the gluteus maximus (−33.5 ± 83%), adductor magnus (−18.4 ± 55.4%) and the vastus muscle groups. All muscle groups showed a similar volume reduction between −0.3% to −2% once corrected for the baseline size. Women showed a more heterogeneous change in muscle volumes, with modest declines in several muscles groups such as the semimembranosus (−3.1 ± 6.8%), the gluteus minimus (−2.6 ± 5.5%) and the vastus muscles (approximately −2%). However, a few smaller muscle groups exhibited an increase in volume over the follow-up period, including the adductor brevis (2.1 ± 16.4%), the pectineus (1.4 ± 9.6%), the sartorius (1.5 ± 6.3%) and the pelvic floor (0.5 ± 4.3%). While men overall showed a more consistent muscle loss across the hip and thigh muscles, they also showed greater individual variability in longitudinal muscle change, as indicated by the higher standard deviations of percentage change.

As well as reductions in individual muscle volumes, we observed increases in rFF at follow-up in both men and women (Supplementary Table S4 provides both absolute and relative percentage changes). Increases in rFF were consistently larger in women. The greatest increases were observed in the posterior thigh muscles, including the biceps femoris (men: 6.2 ± 13.3%; women: 8.0 ± 15.4%), the semimembranosus (men: 7.1 ± 14.1%; women: 8.8 ± 15.1%), and the gluteus minimus (men: 4.2 ± 12.2%; women: 8.3 ± 13.2%).

The annualised rate of change in muscle volume was examined across all 20 bilateral muscle groups as a function of BMI and sex. The cohort was first stratified by sex and subsequently subdivided into 10-year age bands and increasing BMI categories. Distinct sex- and BMI-dependent patterns of muscle volume change were observed across multiple muscle groups (Supplementary Figures S4–S23 and Supplementary Table S12). Across BMI groups, annualised changes in thigh and gluteal muscle volumes were generally modest, but several muscles showed clear evidence of decline. There were sex differences, with some muscles showing significant changes in only one sex. The steepest declines were seen in the adductor longus (− 0.31, *p* = 0.011, in women), biceps femoris (− 0.14, *p* = 0.027, in men), gluteus maximus (−0.35, *p* = 0.011, in women), semimembranosus ( −0.18, *p* = 0.003, in men) and vastus medialis (−0.19, *p* = 0.04, in women). In general, women with a BMI in 10-20 kg/m^2^ consistently showed some of the steepest declines in the cohort, with the changes in eight muscles achieving statistical significance. Interestingly, men showed the steepest declines in muscle volume across the mid-range of BMI (20-30 kg/m^2^), reaching statistical significance in all three adductor muscle groups, the biceps femoris, gluteus muscles, semimembranosus, short external rotators, vastus intermedius, and vastus lateralis. The gluteus muscles consistently showed the greatest statistically significant declines across all BMI groups in both sexes with age. In contrast, neither the gracilis nor the pelvic floor muscles showed any significant changes with age across BMI groups and gender.

### The Influence of type-2 diabetes on muscle volume and relative fat fraction

We matched 1,716 participants with T2D in a case-control study to determine whether generalised changes in muscle volume and quality could be observed for muscle groups in the hips and thighs. The demographics of this cohort are described in Supplementary Table S5. Despite the groups being similar in terms of age, weight, height and BMI, VAT volume was substantially higher in participants with T2D.

When analysed by sex, differences related to T2D status were generally more pronounced in men than in women (Supplementary Table S6). In women, muscle volumes were broadly similar between most muscle groups. Modest, but not statistically significant, reductions were observed in the adductor longus (T2D: 255 ± 53 ml; controls: 264 ± 51 ml), pelvic floor (T2D:117 ± 16 ml; controls 120 ± 15 ml) and rectus femoris (T2D: 322 ± 55 ml; controls: 334 ± 56 ml), while a small increase was observed in the adductor brevis (T2D: 108 ± 24 ml; controls: 105 ± 22 ml). In contrast, men with T2D exhibited consistently lower muscle volumes across the majority of hip and thigh muscles. Larger differences were observed in major muscle groups, including the gluteus maximus (T2D: 1, 897 ± 356 ml; controls: 1, 969 ± 337 ml), rectus femoris (T2D: 442 ± 77 ml; controls: 475 ± 80 ml), and vastus lateralis (T2D: 1, 066 ± 181 ml; controls: 1, 118 ± 173 ml). Smaller differences were observed in several smaller muscles for men, including the piriformis and short external rotators.

Sex-differences were also found in fat accumulation in participants with T2D and their case-controls (Supplementary Table S7). In women, rFF was comparable between most muscle groups. Significant increases were observed in two muscles: the gluteus maximus and gluteus medius. Modest, but not statistically significant, reductions were found in the adductor longus, tensor fasciae latae, vastus intermedius, and vastus medialis. In contrast, men with T2D exhibited consistently higher rFF across almost all muscle groups compared with controls. Larger differences were observed in several hip and thigh muscles, including the gluteus maximus (T2D: 20.9 ± 10.0%; controls: 18.1 ± 8.8%), tensor fasciae latae (T2D: 19.1 ± 9.5%; controls: 17.4 ± 8.5%), and semimembranosus (T2D: 14.0 ± 7.8%; controls: 12.5 ± 6.6%).

### The influence of GLP-1RA on muscle volume and relative fat fraction

Thirty eight participants who had been prescribed GLP-1RA for more than six months were matched with controls. The demographics of this cohort are described in Supplementary Table S8. Overall, there were very few demographic differences between subjects taking GLP-1RA and the matched controls. Only modest differences in absolute muscle volumes were observed in the GLP-1RA and case-control cohorts, with no muscle groups reaching statistical significance after multiple comparisons correction (Supplementary Table S9). Muscle rFF values were consistently lower in the GLP-1RA treated cohort compared to controls in women, whereas the opposite pattern was observed in men. However, none of the differences reached statistical significance after correcting for multiple comparisons (Supplementary Table S10).

## Discussion

We present an automated 3D method for segmenting hip and thigh muscles, enabling robust quantification of both volume and rFF across 20 individual muscles in each leg. Applying these methods at scale allows variation related to biological sex, ageing, metabolic disease, medication use, and asymmetry to be characterised with greater precision than compartment-level approaches. The UK Biobank rescanning cohort further enables examination of short-term longitudinal change, revealing distinct sex-specific patterns of muscle loss and alterations in rFF over an interval of two and a half years.

Current imaging assessments of muscle health rely on broad volumetric measures or coarse compartmental divisions, largely because manually segmenting individual muscles from images is prohibitively time-consuming^26^. While deep learning enables large-scale quantification of muscle volumes, automated methods differ markedly in the number and type of muscles they can segment. A wide array of 2D, 3D, hybrid, and multi-atlas architectures has emerged, offering important advances but highlighting substantial inconsistency in anatomical coverage, data requirements, and performance^25,28,28–31,31–40^. The literature increasingly demonstrates the feasibility and fragmentation of automated muscle segmentation, underscoring the need for scalable, standardised models that reliably capture fine-grained muscle-level heterogeneity. Dixon MRI additionally enables estimates of rFF, a clinically-relevant biomarker given the strong links between muscle fat accumulation and metabolic disease.

In agreement with Lin *et al*.^36^ and Kim *et al*.^40^, our model performance measured by Dice score was strongly related to muscle volume, with larger muscles like the gluteus maximus performing better (0.940) than smaller muscles such as the adductor brevis and pectineus, although only these two muscles had a Dice score < 0.8, while nine muscles had a Dice score > 0.9. These results indicate that while segmentation accuracy naturally scales with muscle size, our model delivers consistently high performance across the vast majority of muscle groups. Our analysis enabled a clear assessment of left–right symmetry in muscle volumes at the cohort level, with greater variability in the smaller muscle groups. This approach was also useful for identifying pronounced, highly localised asymmetries in individuals, which differentiates pathological atrophy from normal anatomical variation.

The rank order of individual muscle volumes was consistent between men and women, with the largest muscles (gluteus maximus, adductor magnus, vastus lateralis, vastus intermedius and vastus medialis) and the smallest muscles (adductor brevis, pelvic floor, pectineus, short external rotators and piriformis) following the same descending pattern in both. Men had higher muscle volumes than women overall and for each muscle group measured, although the magnitude of the sex differences was reduced after height normalisation. Volumes were comparable in smaller muscles such as the short external rotators, piriformis, and pectineus. Women had consistently higher rFF than men across all muscle groups. In men, rFF was highest in the gluteus maximus, whereas in women, it was highest in the tensor fasciae latae; however, the same five muscles ranked in both sexes.

Sex differences in muscle fat content are inconsistently reported, with studies showing higher levels in men^53^, no differences^54^, or higher levels in women^55^. Muscle fat is influenced by fibre type, oxidative Type I muscles typically show higher fat than glycolytic Type II^56^, and anterior thigh muscles generally have lower fat than posterior muscles^57^, reflecting the greater abundance of Type I fibres posteriorly^58^. Men also tend to have larger fibres and a higher proportional area of Type II fibres^20^. However, these relationships did not consistently align with rFF patterns in our data. For example, the tensor fasciae latae, (Type II rich) showed the highest fat content in women, whereas the Type I rich vastus intermedius and vastus medialis, had the lowest fat in both sexes. Overall, rFF was higher in postural and pelvic stabilising muscles around the hip, (e.g., gluteus maximus, tensor fasciae latae, sartorius); and lower in load-bearing and force-generating muscles of the thigh and knee.

Integrating muscle imaging with histology is challenging since studies differ in how they quantify muscle fat and in which fat depot they measure. Muscle fibre type and size vary with sex, ageing, disuse, training, and disease^20^ and most histological studies examine only small samples and few muscles. Many MRI/CT studies assess intramuscular adipose tissue (e.g., Goutallier grading)^59^, rather than the fat stored within lean muscle tissue, which is the focus here. Additional complexity arises from the range of MRI techniques used to quantify intermuscular and intramuscular fat, PDFF, and rFF, while ^1^H-MR spectroscopy uniquely distinguishes intra-from extra-myocellular lipids. This is something conventional MRI cannot yet achieve.

Across two and a half years, UK Biobank participants showed a clear shift toward a less favourable body-composition profile. In men, most individual muscles exhibited small but consistent volume losses, whereas women showed a more heterogeneous pattern, with several smaller muscles demonstrating slight increases. Although mean changes were modest (approximately ± 2%, where all percentage changes are in Supplementary Tables S3 and S4), the absolute losses were greatest in the largest muscles. When expressed relative to baseline size, however, the pattern was more uniform: for example, the gluteus maximus declined by approximately 33 ml in men, yet this represented only a 1.7% reduction, comparable to the proportional changes observed in much smaller muscles. Annualised changes also varied by BMI and sex, with alterations in specific BMI groups. The finding that women with low BMI showed clear declines in multiple muscle groups is notable and may suggest a particular risk in this population. Taken together, these results highlight which muscles are most sensitive to ageing in mid-life and early older age.

Increases in rFF accompanied changes in volume, particularly in women, potentially reflecting higher sedentary time at follow-up. Preferential atrophy of Type II fibres^58^ may also contribute to a higher-fat phenotype, although a two-year interval may be too short for this to fully emerge. Overall, our findings align with recognised features of sarcopenia, declines in both muscle mass and quality, suggesting that MRI-derived markers may detect early metabolic deterioration within muscle, before overt contractile deficits are apparent. This may be relevant for rehabilitation.

In participants with T2D, we observed marked sex differences in muscle composition; men showed lower muscle volumes and higher rFF across almost all muscles. This is consistent with previous reports of reduced muscle mass^60^, accelerated age-related atrophy^61^, and metabolic disturbances of T2D, which reduce muscle regenerative capacity^62^. In contrast, women with T2D showed minimal differences in muscle volume compared with controls, and heterogeneous rFF changes, with increases in a small minority of muscles. This pattern aligns with evidence that women are generally more resistant to muscle loss, partly due to oestrogen’s protective effects on mitochondrial function, inflammation, and muscle protein synthesis^63^. However, this advantage is diminished in T2D and further reduced after menopause, when oestrogen levels decline and metabolic risk increases. In GLP-1RA–treated participants, small, non-significant volume reductions were seen, but limited sample size and missing treatment-timing data prevent firm conclusions.

This study has several limitations. The UK Biobank imaging protocol covers only the neck to the knee region, excluding the lower legs; so our models were restricted to hip and thigh musculature. Very tall individuals with incomplete coverage or those with knee/hip implants were also excluded. Only Dixon MRI is available for the thighs, limiting fat quantification to rFF, which is sensitive to T1/T2 effects, and does not model the fat–water spectrum making it semi-quantitative compared with PDFF. Finally, the quality of segmentation performance scaled with muscle volume; therefore, changes in the smallest muscles should be interpreted cautiously.

This work demonstrates how automated, muscle-level MRI analysis can uncover ageing, metabolic, sex-specific, and treatment-related patterns that remain invisible to compartment-based approaches. By enabling scalable, precise quantification of both volume and fat infiltration across 20 muscles, it establishes a foundation for population-scale musculoskeletal phenotyping. The UK Biobank rescanning data demonstrate that subtle but clinically-meaningful changes in muscle health can be detected over only two years, underscoring the potential of imaging-derived biomarkers to identify early metabolic deterioration before functional decline. Integrating these methods with longitudinal, genetic, and interventional studies will be crucial for mapping individual trajectories of muscle ageing, refining risk prediction for sarcopenia and metabolic disease, and informing personalised prevention and rehabilitation strategies.

## Acknowledgements

This study was carried out using UK Biobank Application number 23889. We thank the participants in the UK Biobank imaging study.

## Author contributions statement

JDB, ELT and BW conceived the study. JDB, BW, ELT, HR and DA designed the study. HR, CBB and DA performed the manual annotations. BW, NB and MT implemented the methods and performed the data analysis. MT defined the disease and physiological condition categories. MT performed the statistical analysis. ELT, BW, MT, MN and DA drafted the manuscript. All authors reviewed the manuscript.

## Data availability statement

Under the standard UK Biobank data sharing agreement, we (and other researchers) cannot directly share raw data obtained or derived from the UK Biobank. Under this agreement all data generated and methodologies used in this paper are returned to the UK Biobank, where they will be fully available. Access is obtained directly from the UK Biobank to all bona fide researchers upon submitting a health-related research proposal to the UK Biobank (www.ukbiobank.ac.uk).

