## Supplementary Figures and Tables for "Heterogeneity, Longitudinal Decline, and Metabolic Risk in MRI-Based Quantification of 20 Individual Hip and Thigh Muscles"

### ABSTRACT

### Supplementary Information

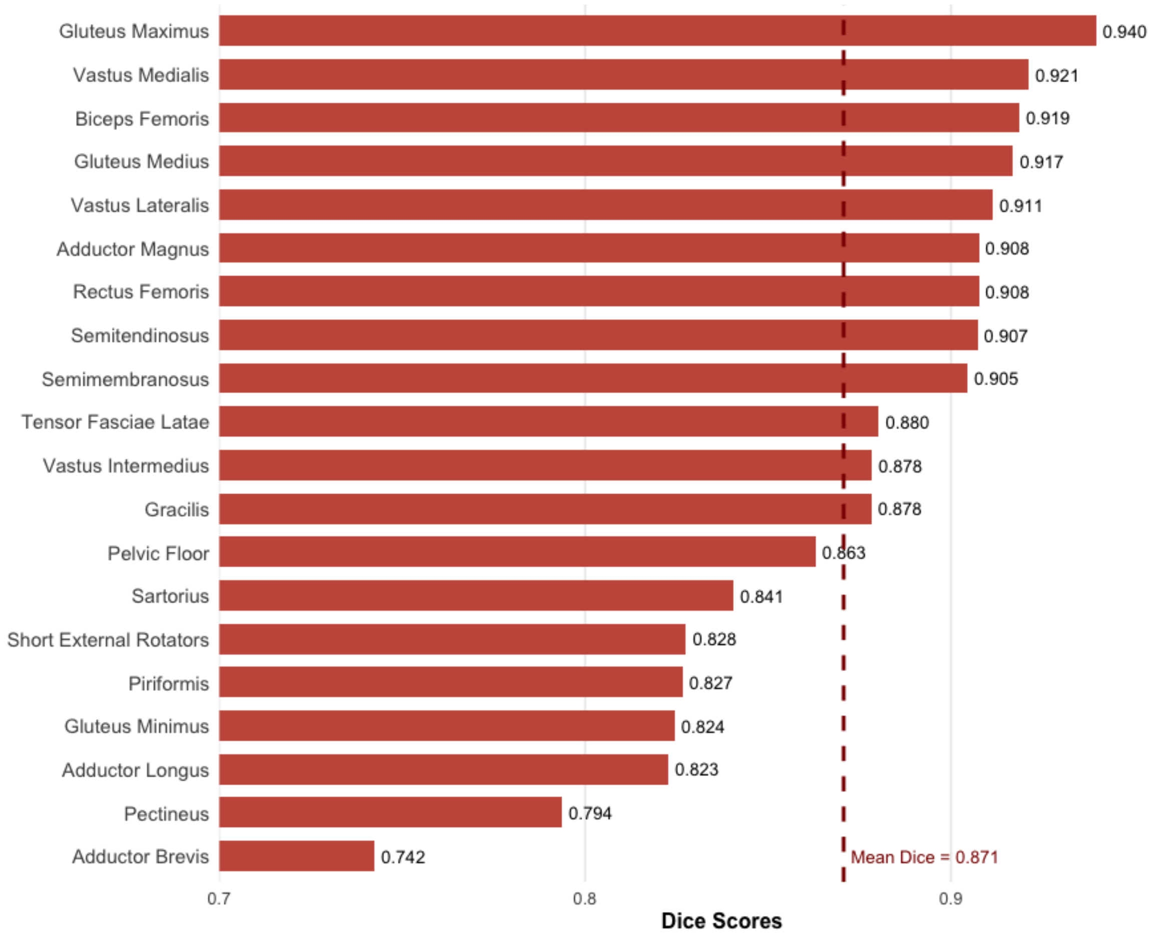


**Figure S1.** Average Dice scores for the segmentation model on 20 bilateral muscle groups in the hips and thighs.

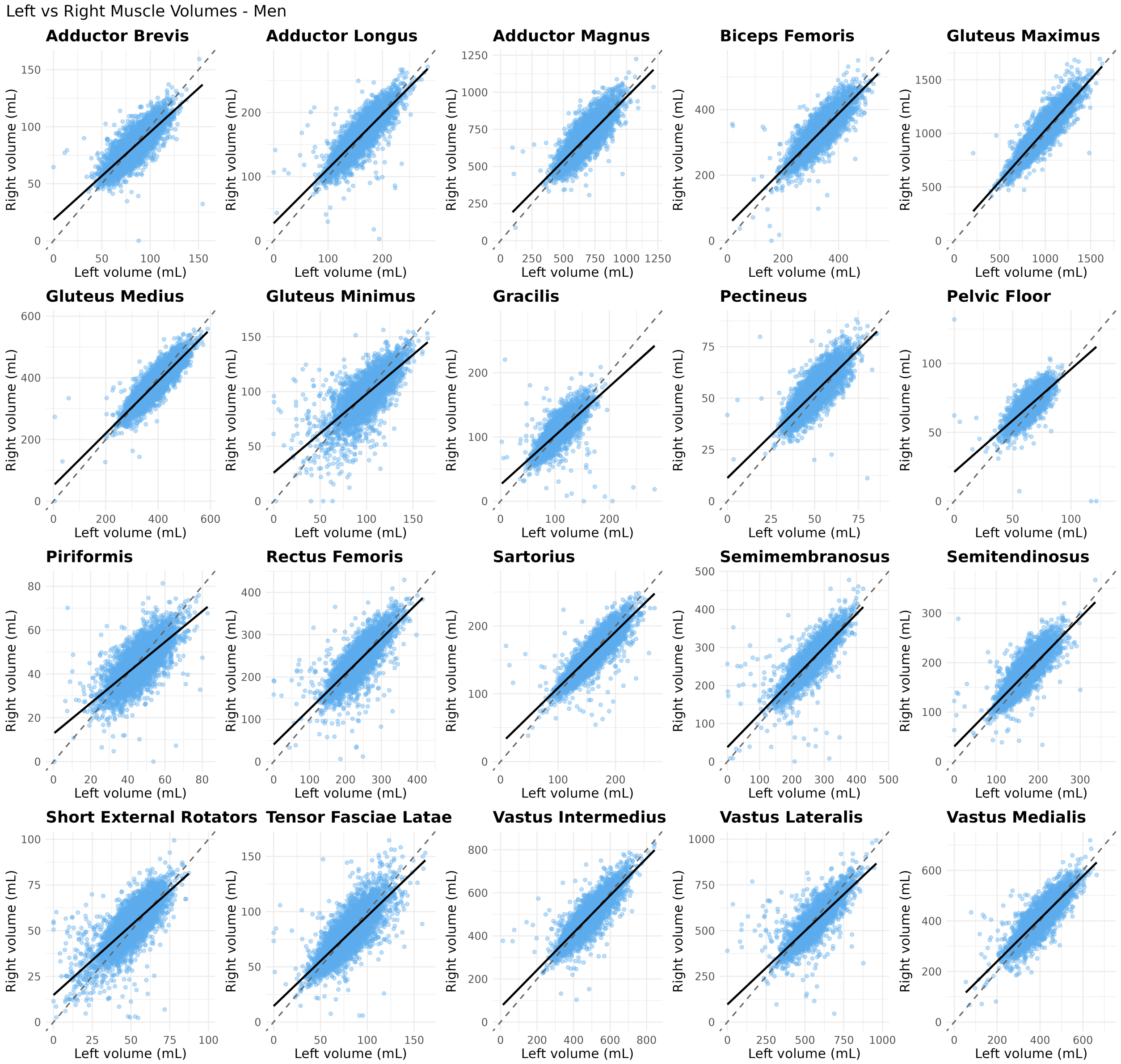


**Figure S2.** Comparison of left and right leg muscle volumes in male participants. Muscle volume in the left leg is plotted on the x-axis and the right leg on the y-axis. The dotted line is the 45° line, denoting equality between the measurements, and the solid line is the linear regression between the measurements.

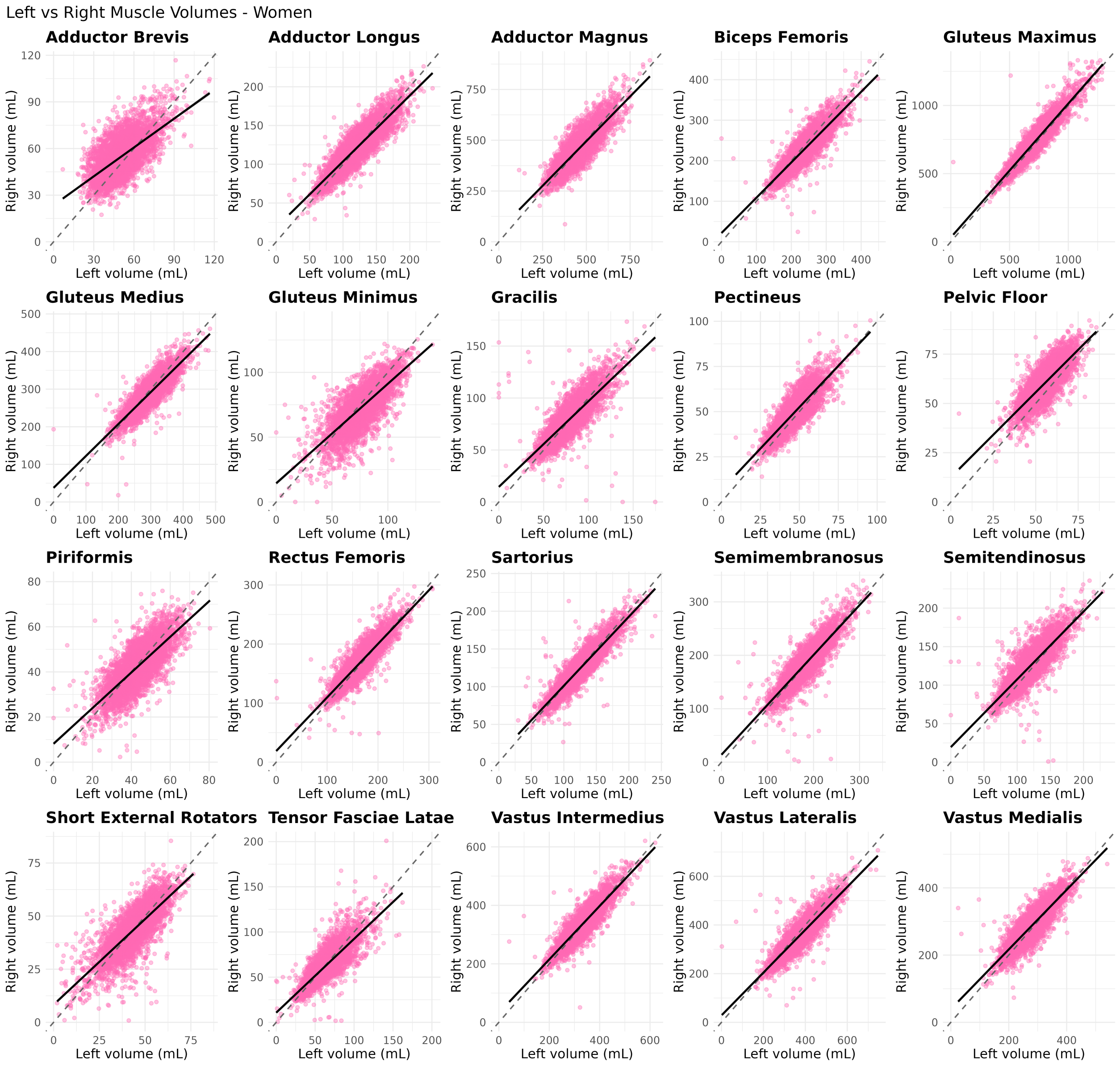


**Figure S3.** Comparison of left and right leg muscle volumes in female participants. Muscle volume in the left leg is plotted on the x-axis and the right leg on the y-axis. The dotted line is the 45° line, denoting equality between the measurements, and the solid line is the linear regression between the measurements.

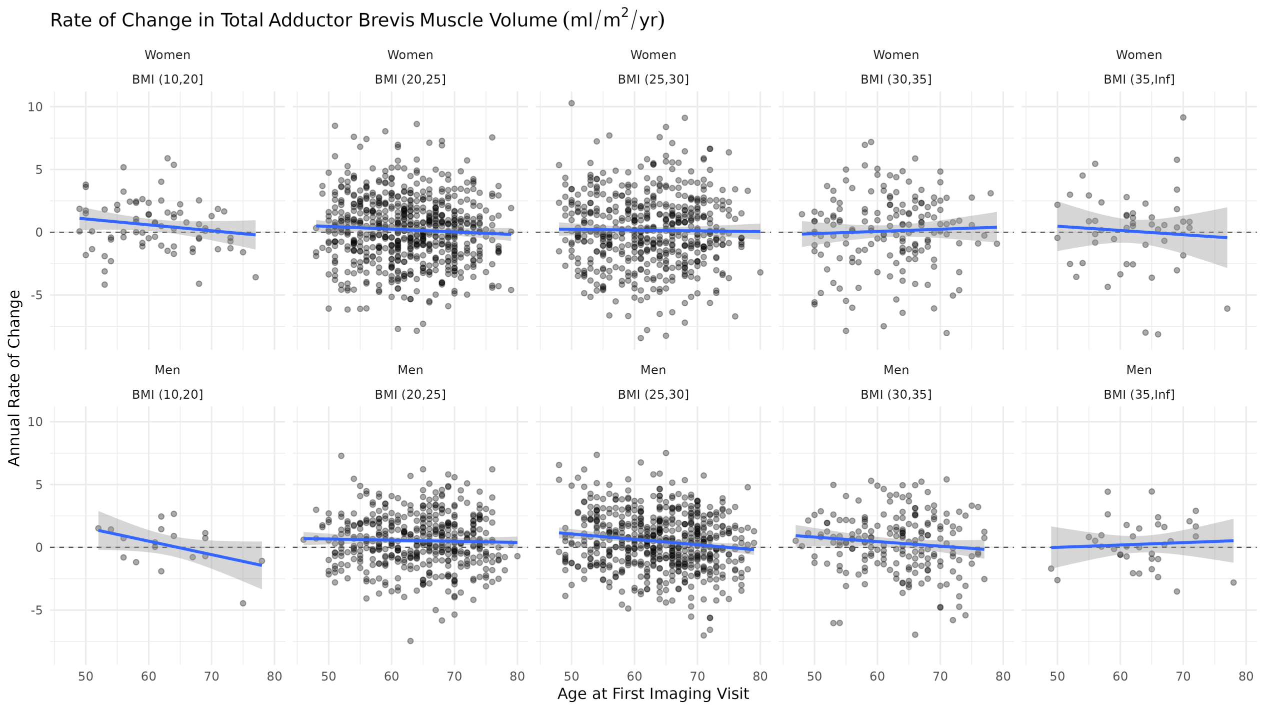


**Figure S4.** Annual rate of change in the adductor brevis muscle from the longitudinal cohort, separated by sex and BMI groups.

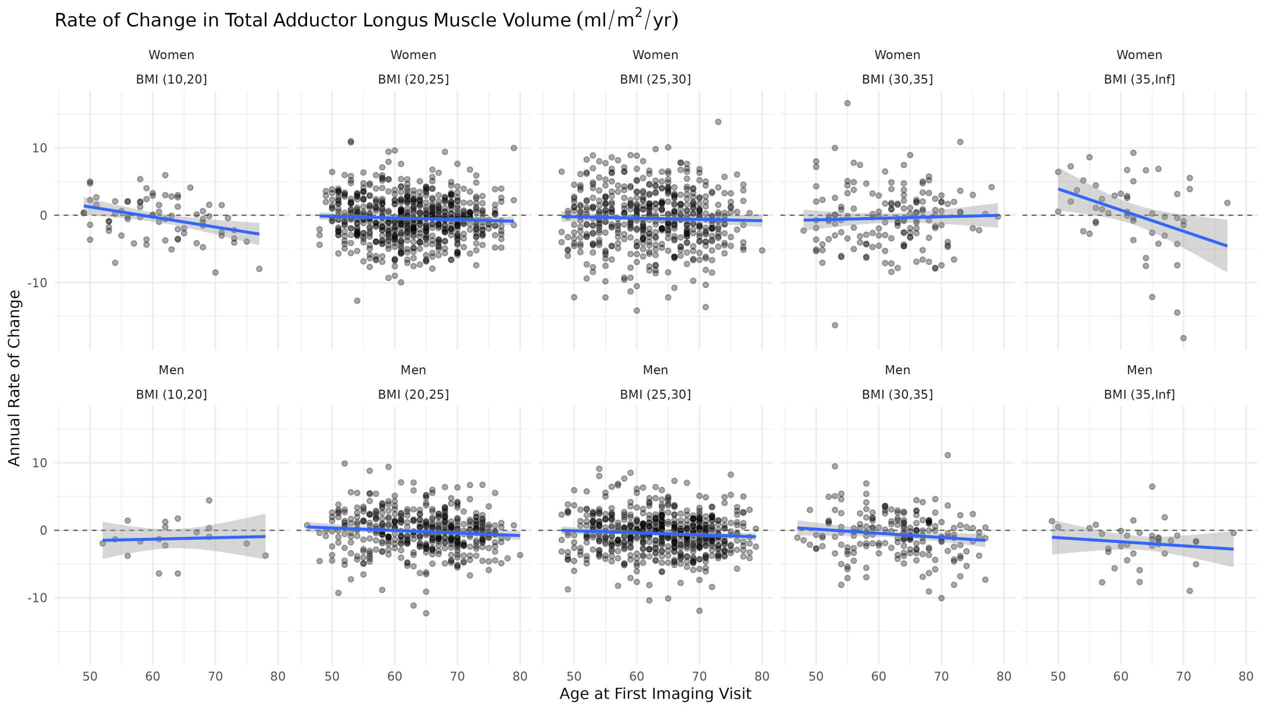


**Figure S5.** Annual rate of change in the adductor longus muscle from the longitudinal cohort, separated by sex and BMI groups.

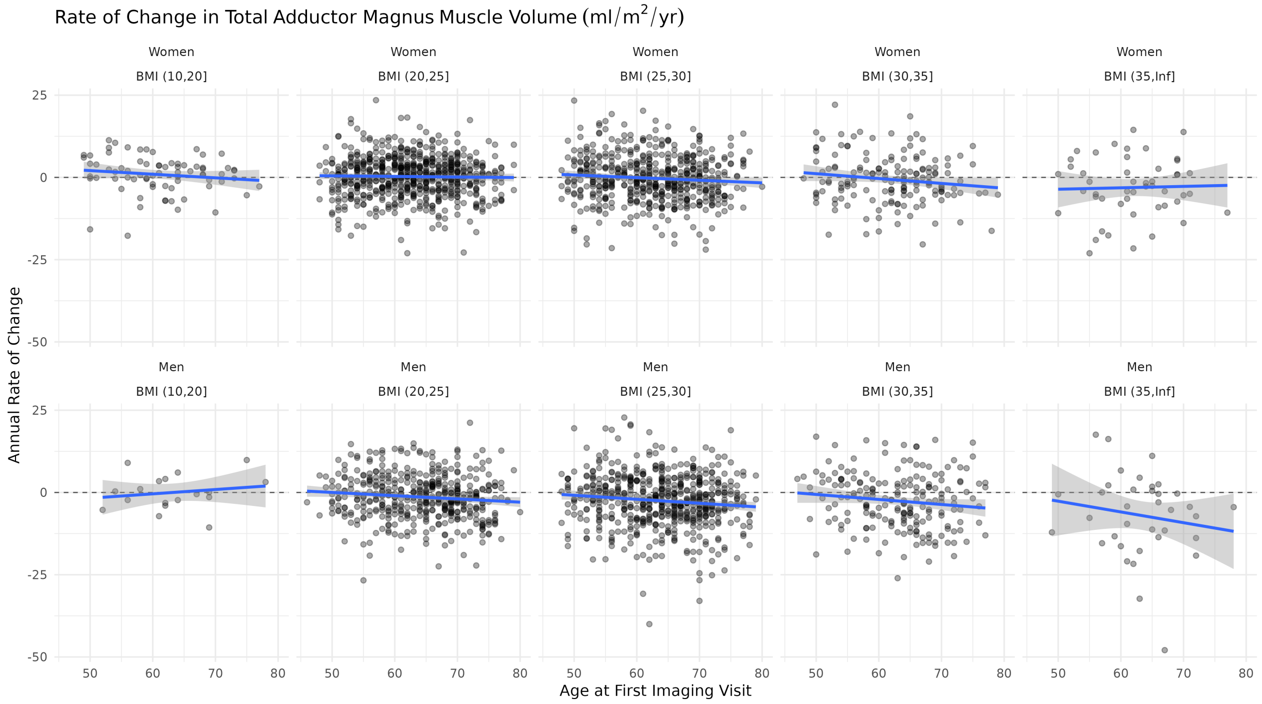


**Figure S6.** Annual rate of change in the adductor magnus muscle from the longitudinal cohort, separated by sex and BMI groups.

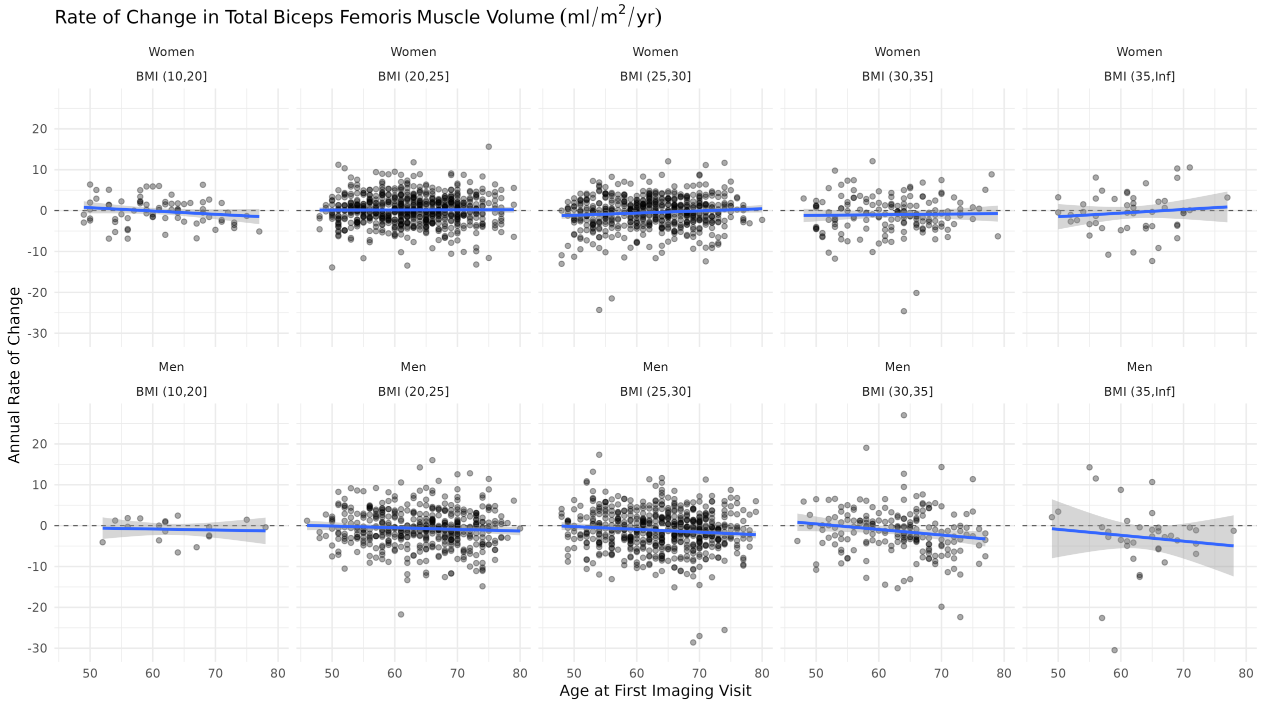


**Figure S7.** Annual rate of change in the biceps femoris muscle from the longitudinal cohort, separated by sex and BMI groups.

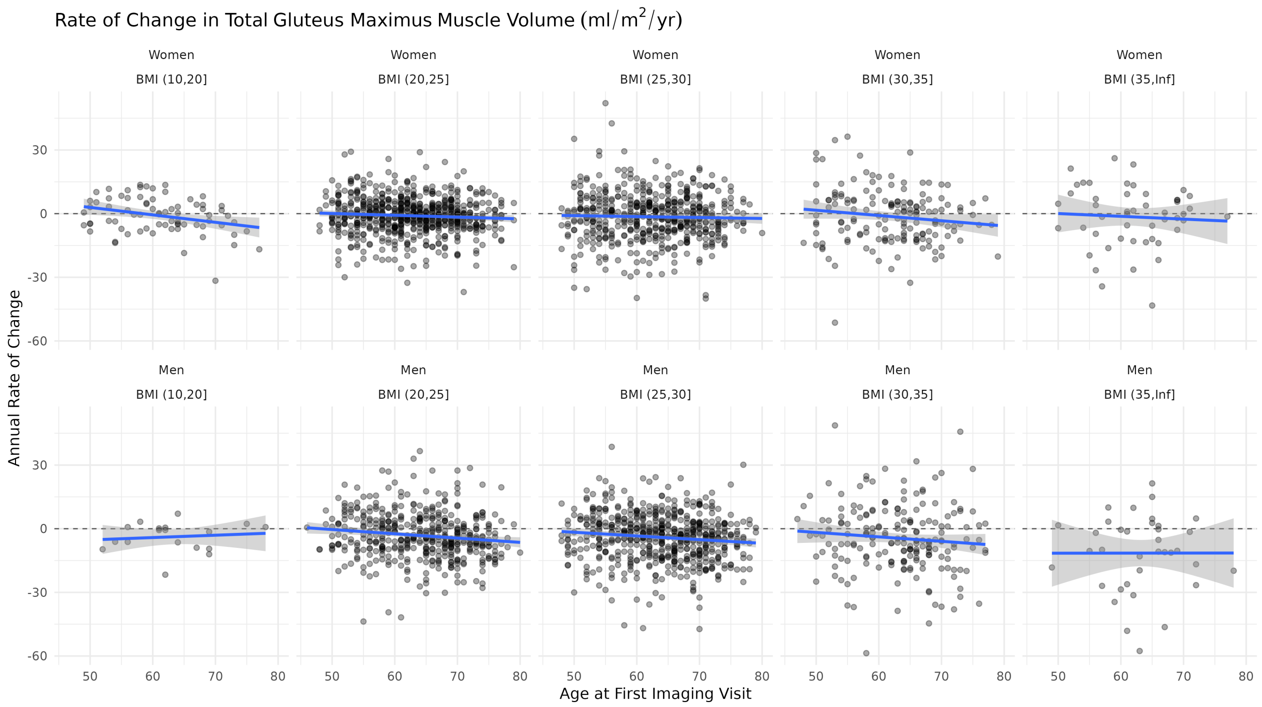


**Figure S8.** Annual rate of change in the gluteus maximus muscle from the longitudinal cohort, separated by sex and BMI groups.

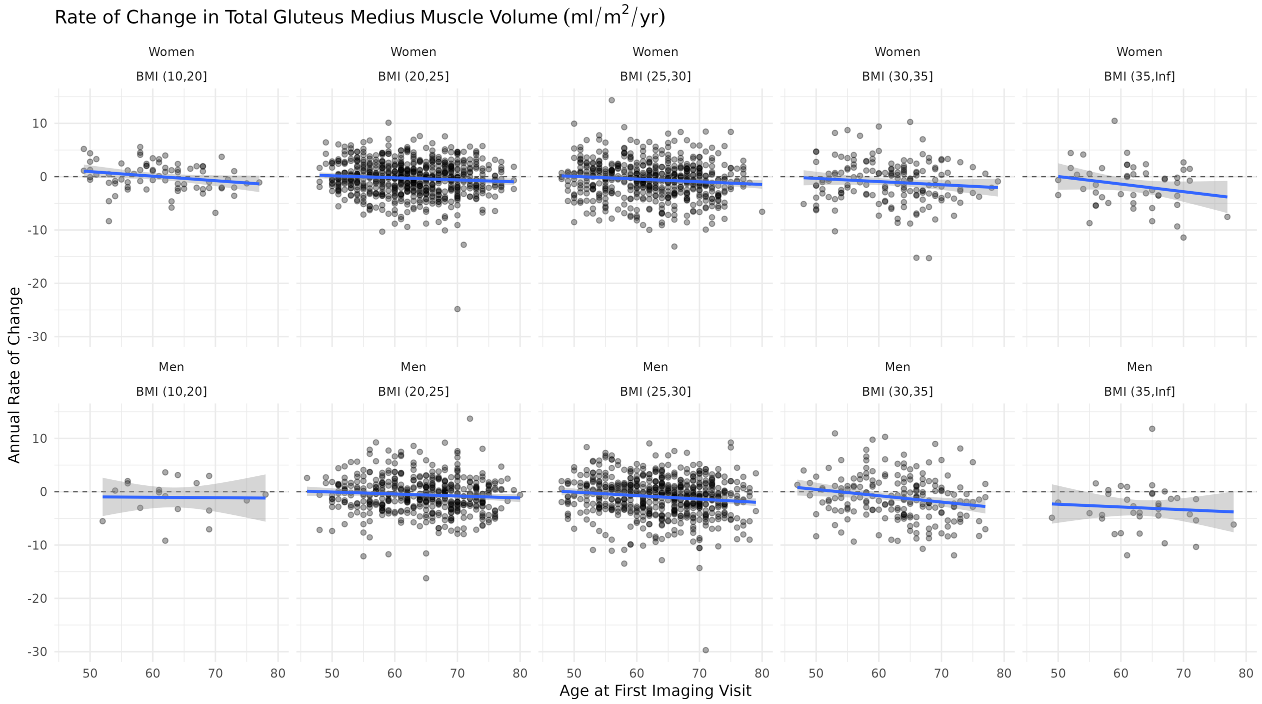


**Figure S9.** Annual rate of change in the gluteus medius muscle from the longitudinal cohort, separated by sex and BMI groups.

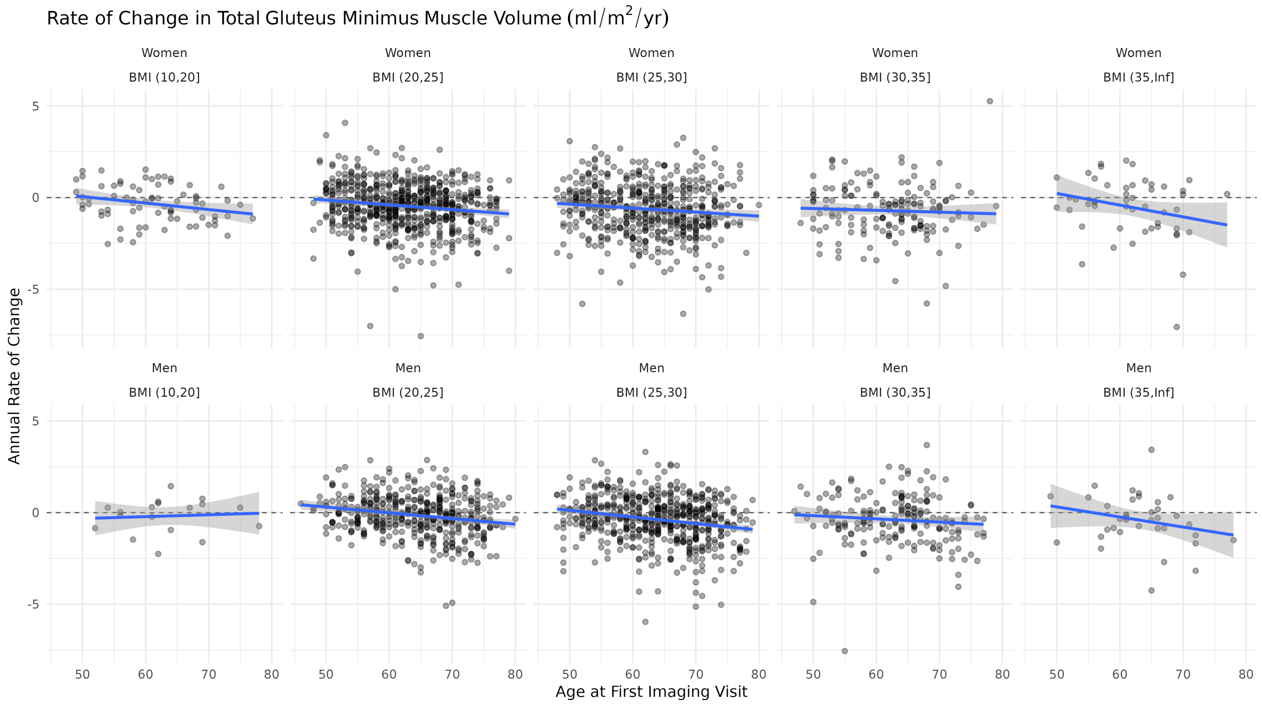


**Figure S10.** Annual rate of change in the gluteus minimus muscle from the longitudinal cohort, separated by sex and BMI groups.

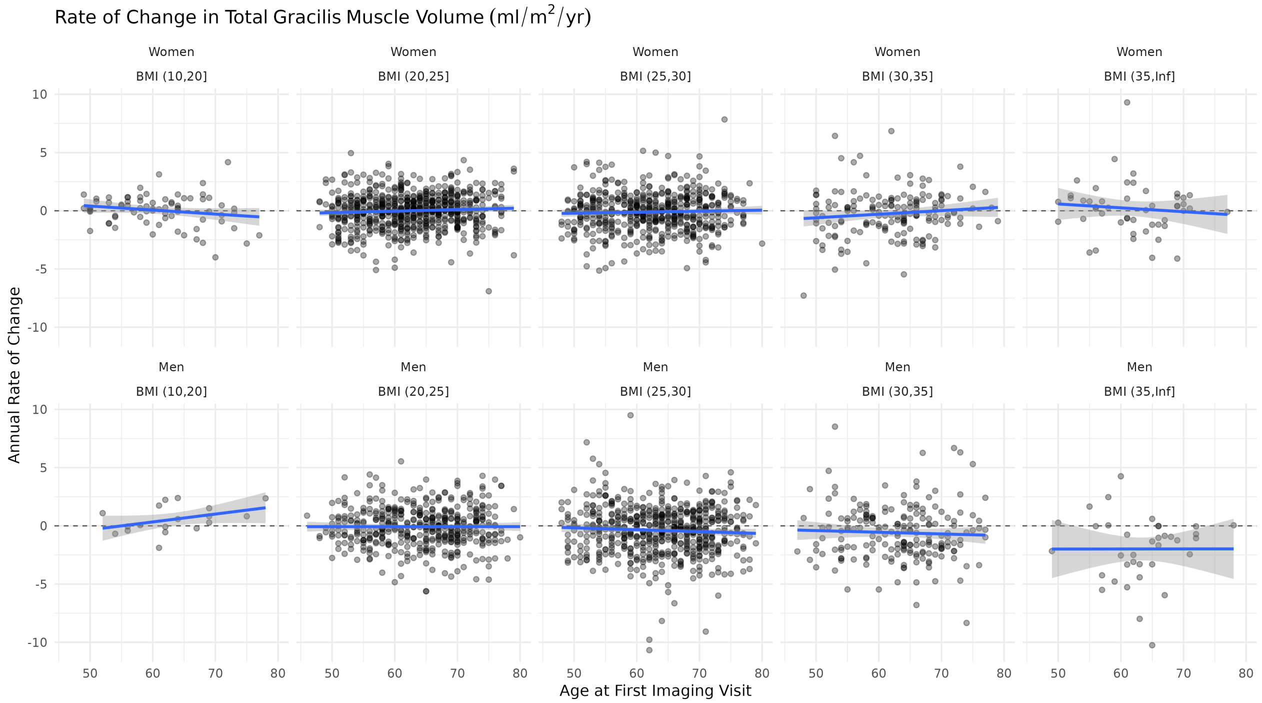


**Figure S11.** Annual rate of change in the gracilis muscle from the longitudinal cohort, separated by sex and BMI groups.

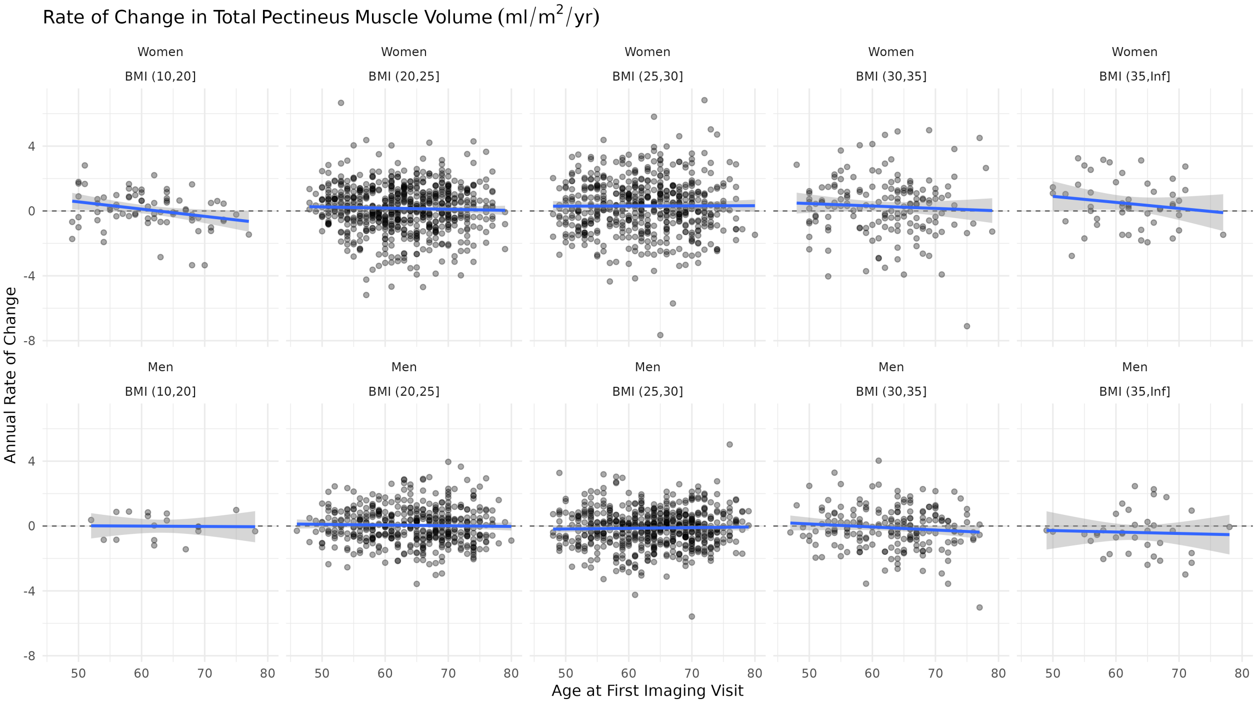


**Figure S12.** Annual rate of change in the pectineus muscle from the longitudinal cohort, separated by sex and BMI groups.

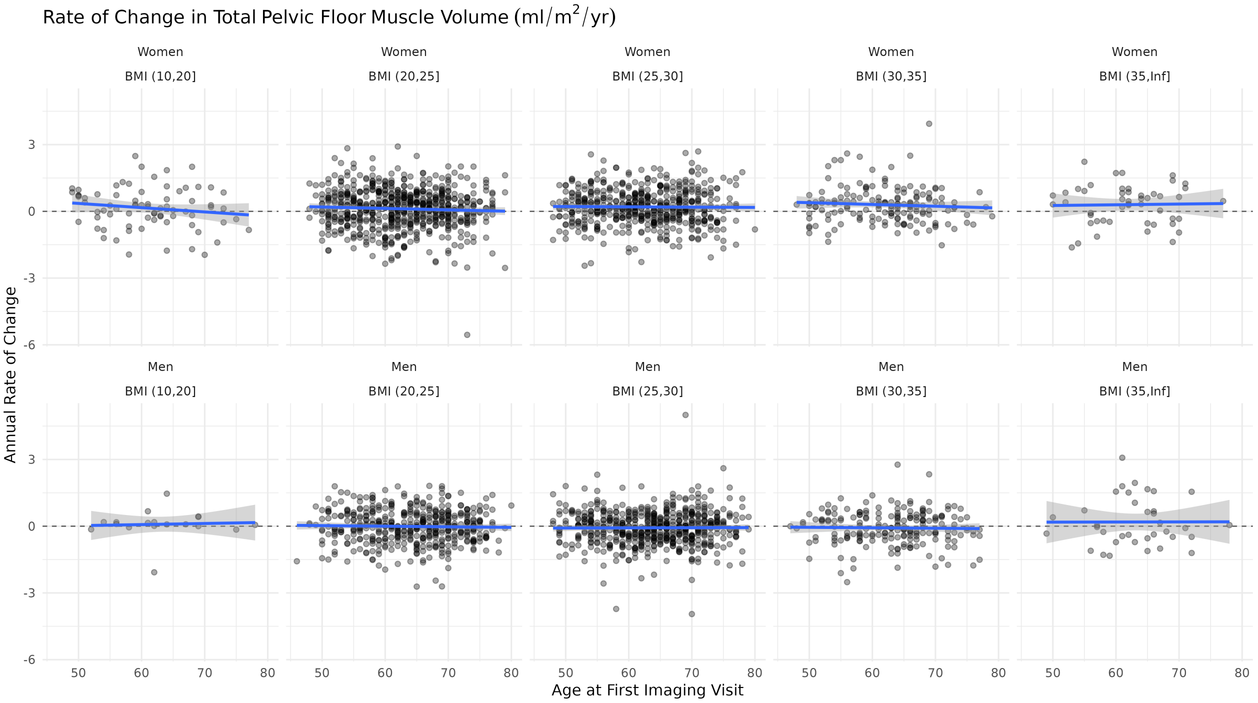


**Figure S13.** Annual rate of change in the pelvic floor muscle from the longitudinal cohort, separated by sex and BMI groups.

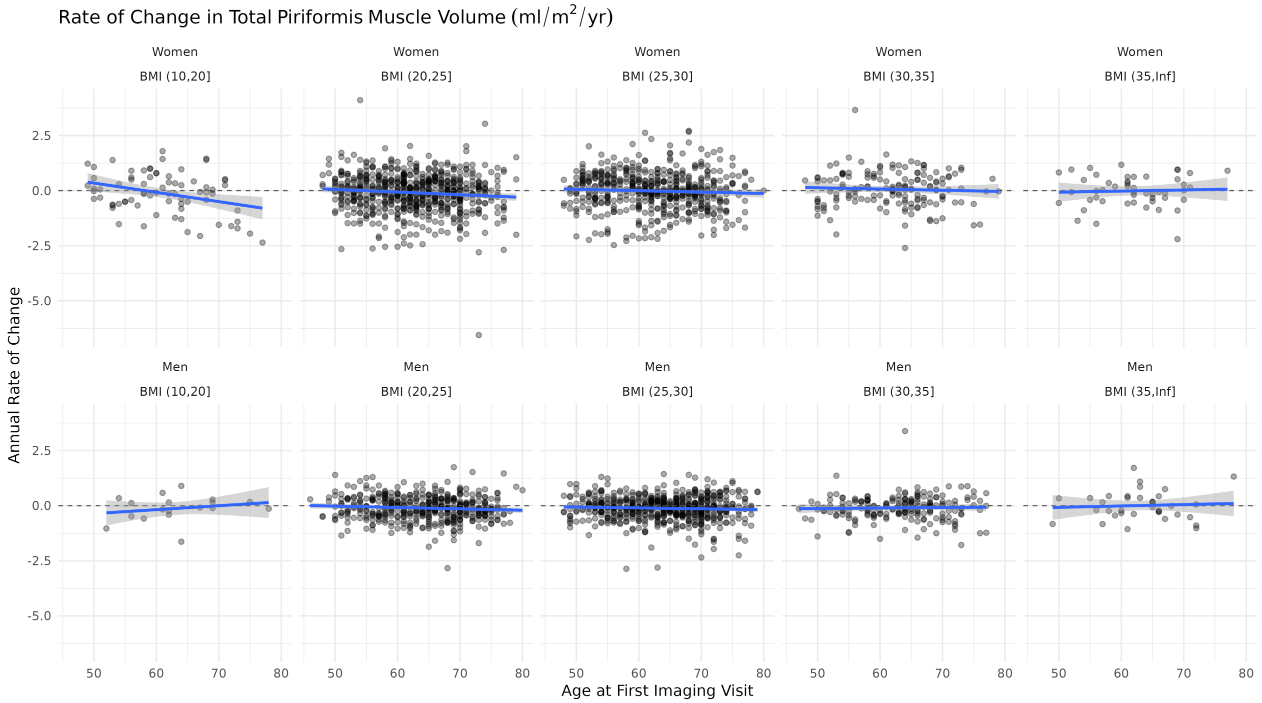


**Figure S14.** Annual rate of change in the piriformis muscle from the longitudinal cohort, separated by sex and BMI groups.

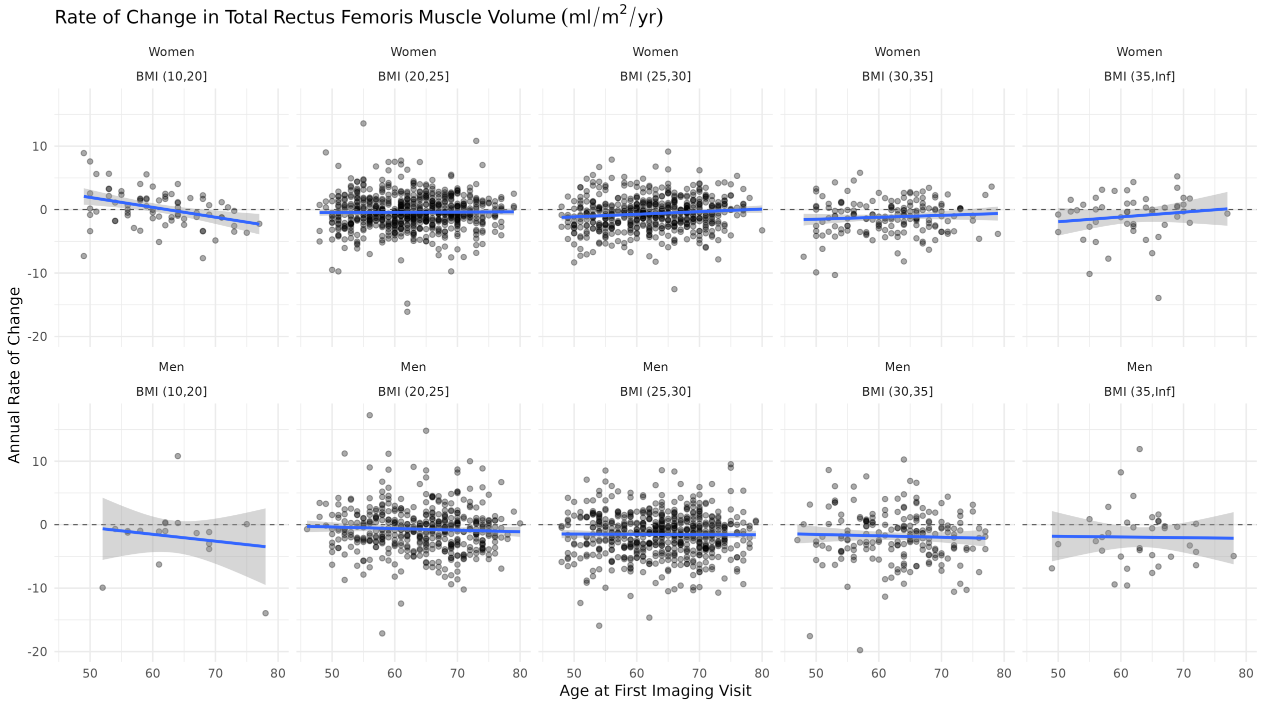


**Figure S15.** Annual rate of change in the rectur femoris muscle from the longitudinal cohort, separated by sex and BMI groups.

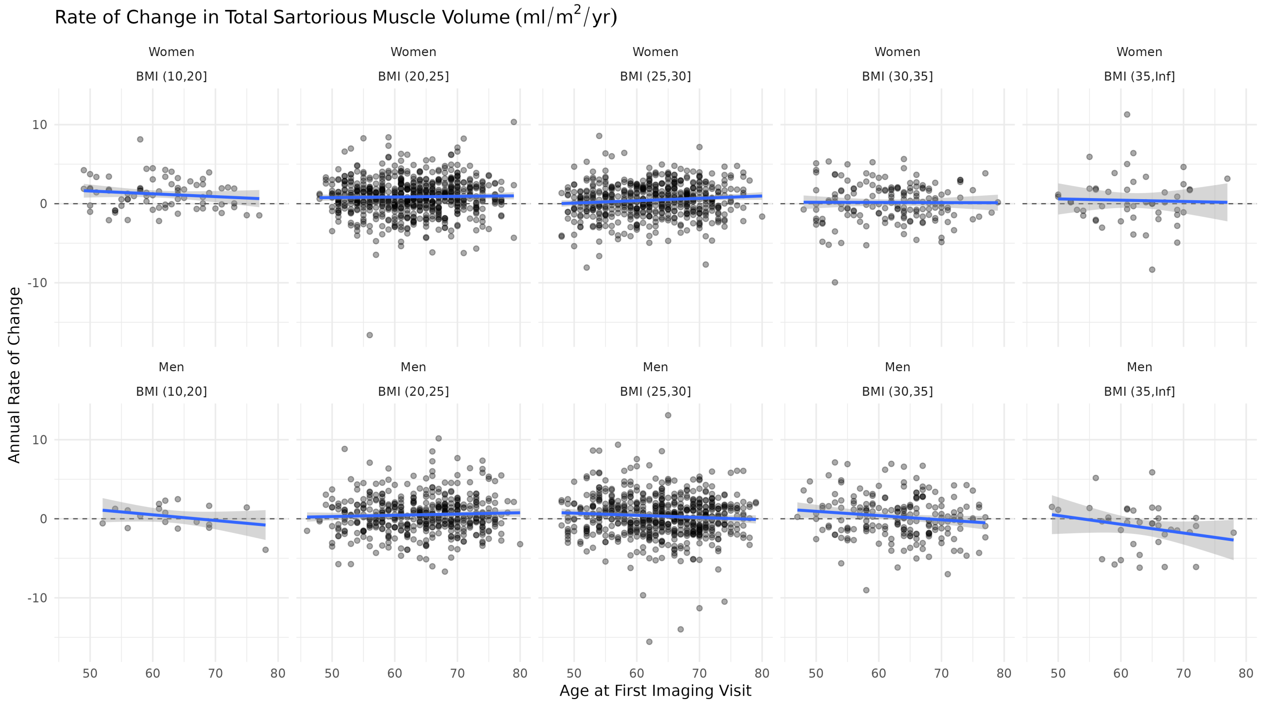


**Figure S16.** Annual rate of change in the sartorius muscle from the longitudinal cohort, separated by sex and BMI groups.

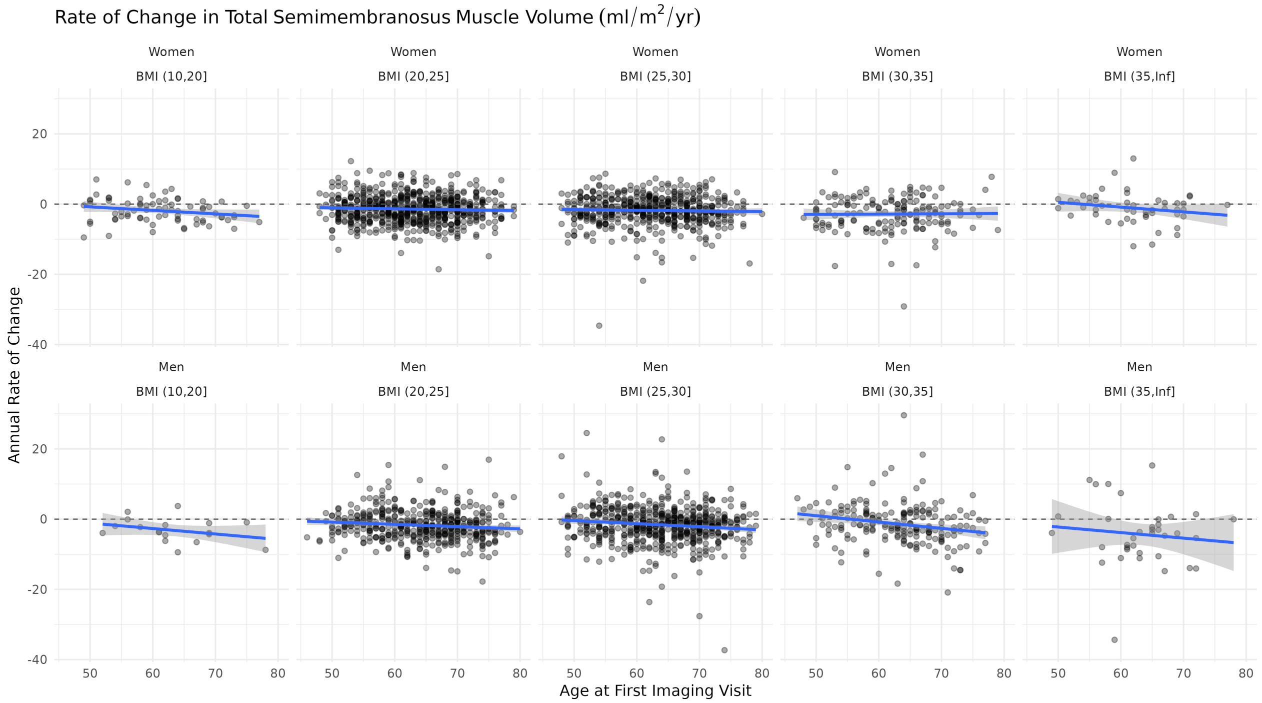


**Figure S17.** Annual rate of change in the semimembranosus muscle from the longitudinal cohort, separated by sex and BMI groups.

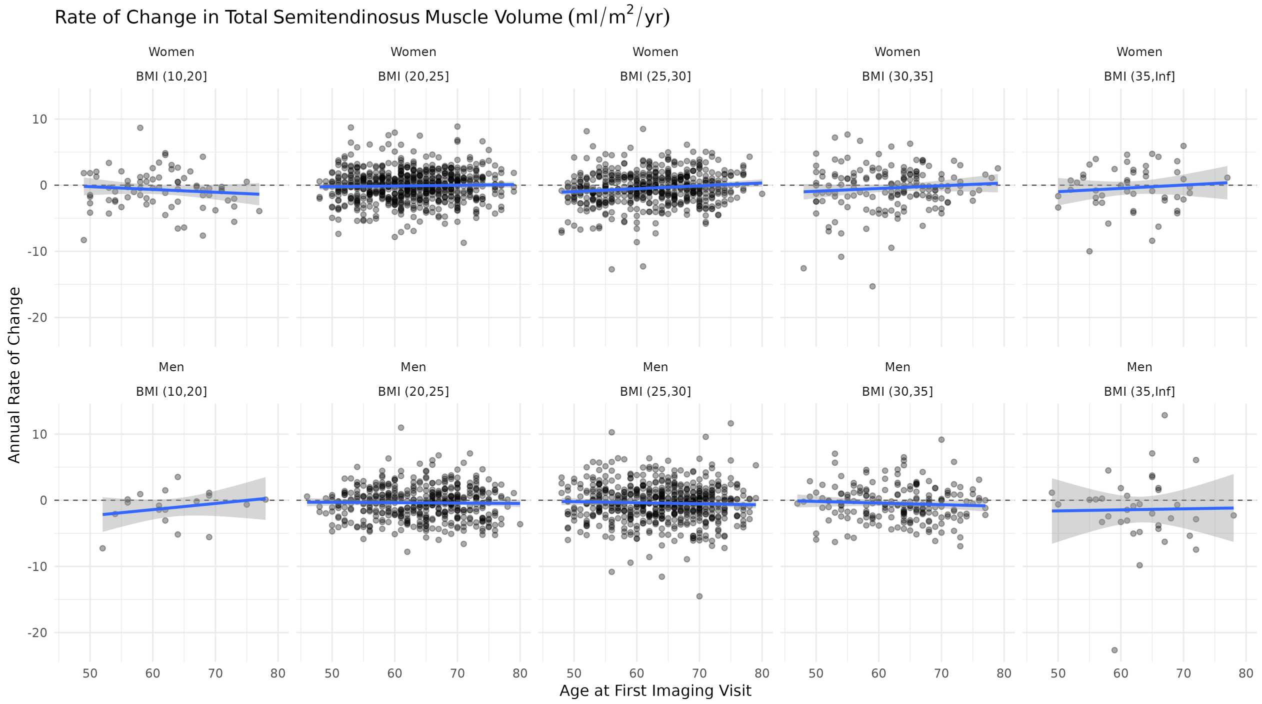


**Figure S18.** Annual rate of change in the semitendinosus muscle from the longitudinal cohort, separated by sex and BMI groups.

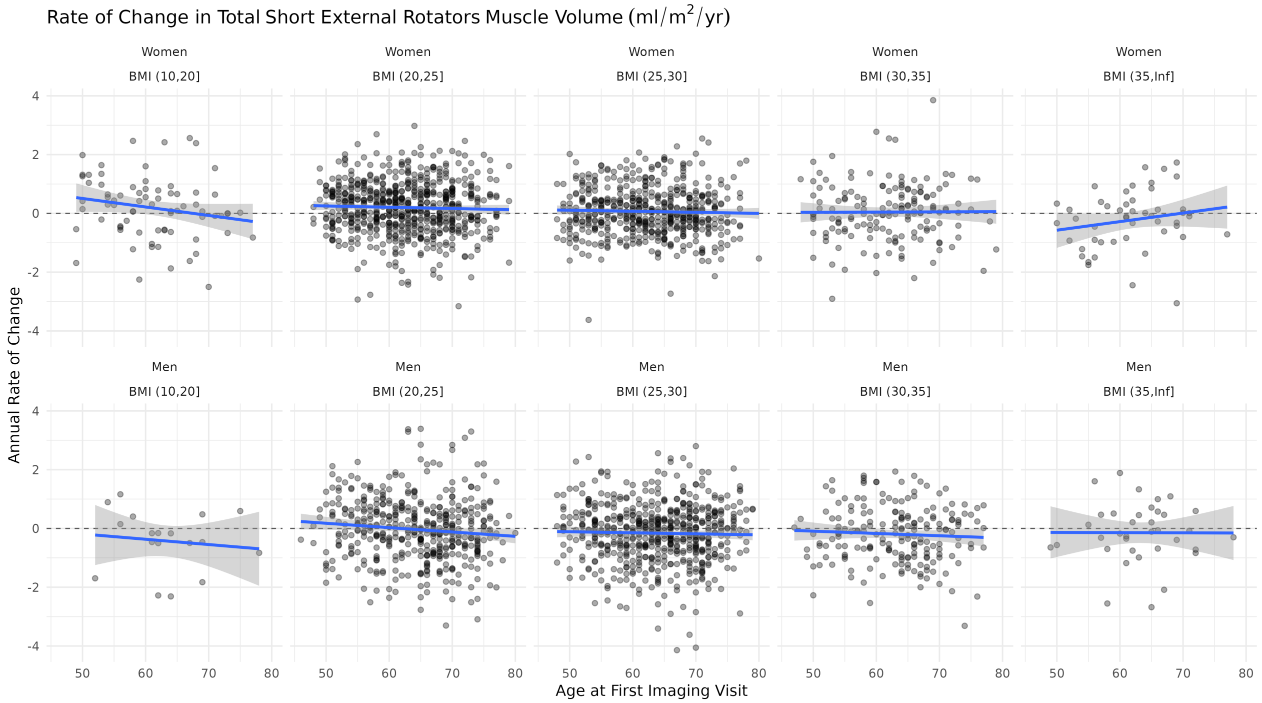


**Figure S19.** Annual rate of change in the short external rotator muscle from the longitudinal cohort, separated by sex and BMI groups.

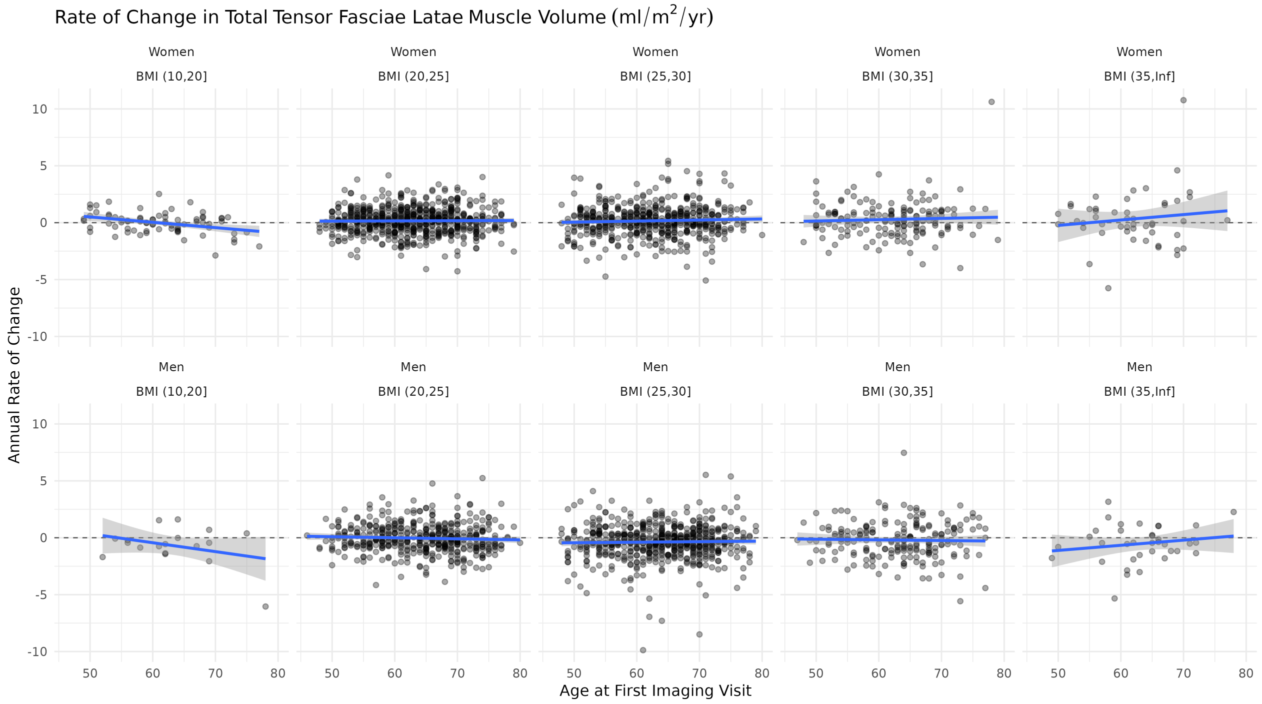


**Figure S20.** Annual rate of change in the tensor fasciae latae muscle from the longitudinal cohort, separated by sex and BMI groups.

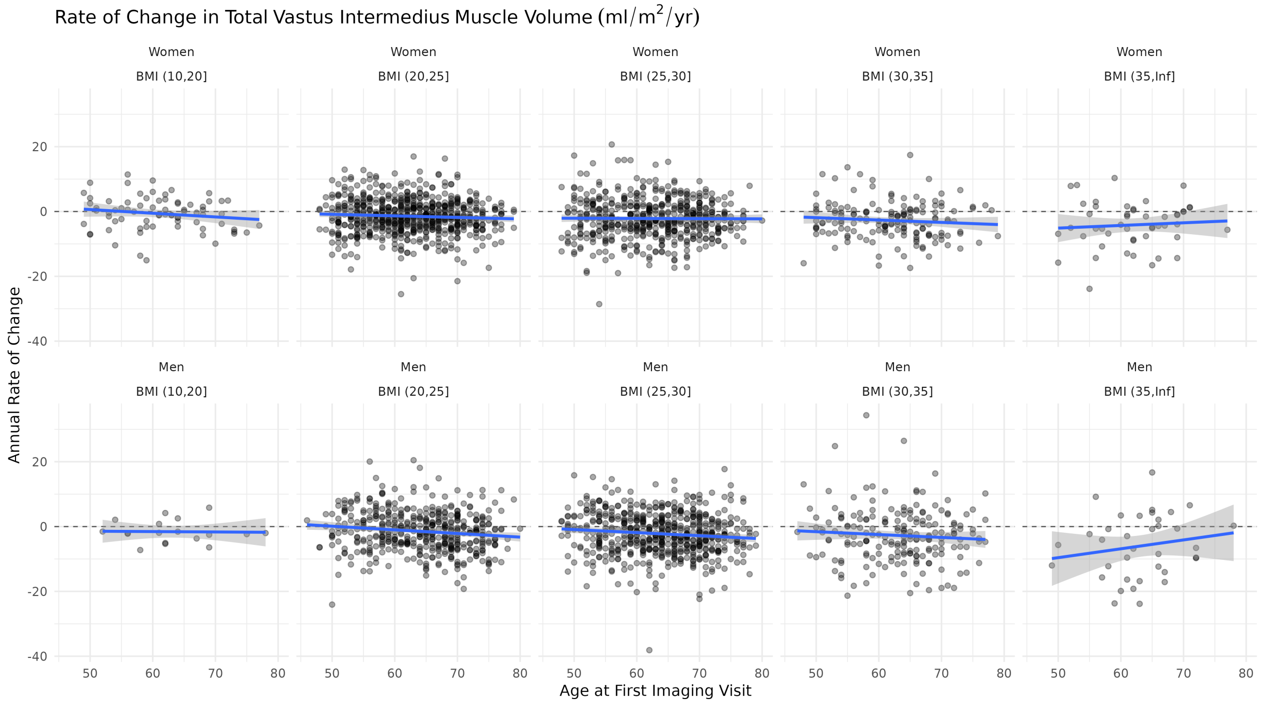


**Figure S21.** Annual rate of change in the vastus intermedius muscle from the longitudinal cohort, separated by sex and BMI groups.

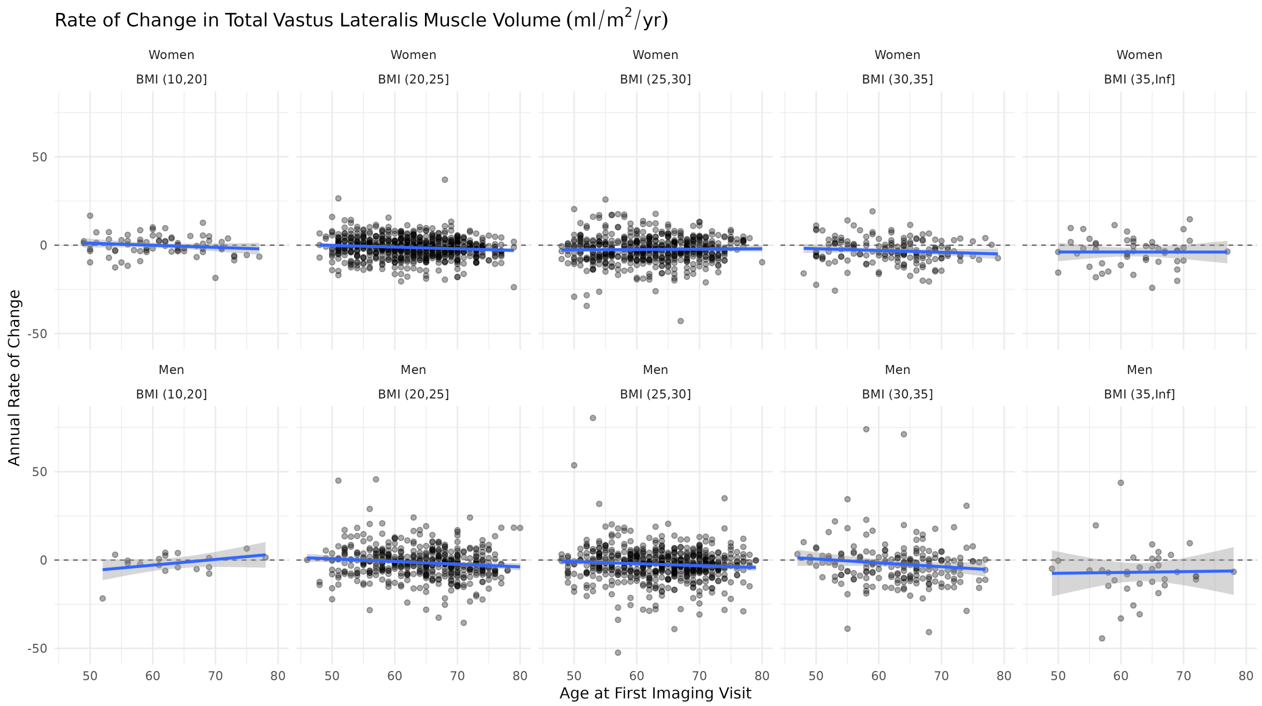


**Figure S22.** Annual rate of change in the vastus lateralis muscle from the longitudinal cohort, separated by sex and BMI groups.

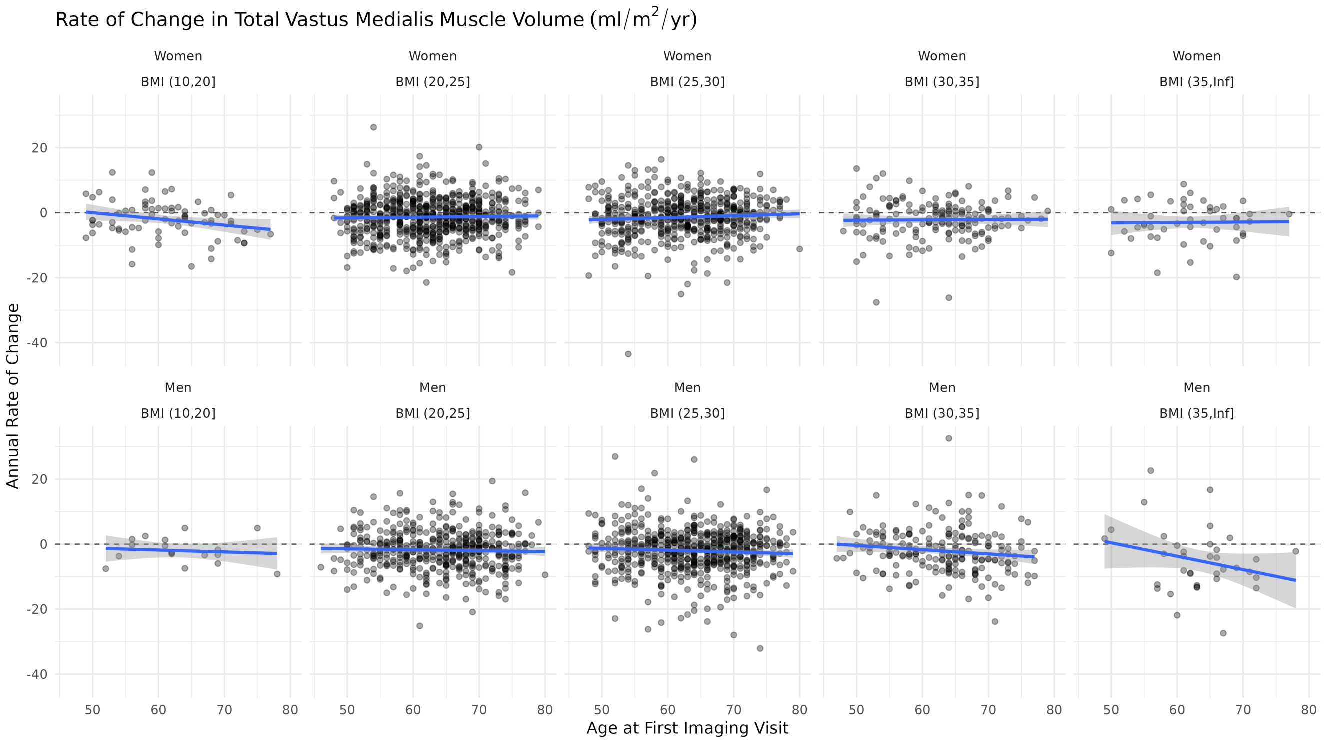


**Figure S23.** Annual rate of change in the vastus medialis muscle from the longitudinal cohort, separated by sex and BMI groups.

| Image-Derived Phenotype | Women<br>$n = 5,199$[1] | Men<br>$n = 5,641$[1] | Overall<br>$n = 10,840$[1] |
|---|---|---|---|
| Adductor Brevis Index ($ml/m^2$) | 40 (8) | 54 (9) | 47 (11) |
| Adductor Longus Index ($ml/m^2$) | 92 (19) | 105 (17) | 99 (19) |
| Adductor Magnus Index ($ml/m^2$) | 341 (57) | 440 (63) | 392 (78) |
| Biceps Femoris Index ($ml/m^2$) | 175 (29) | 214 (31) | 195 (36) |
| Gluteus Maximus Index ($ml/m^2$) | 540 (98) | 625 (101) | 584 (108) |
| Gluteus Medius Index ($ml/m^2$) | 217 (29) | 245 (30) | 232 (33) |
| Gluteus Minimus Index ($ml/m^2$) | 57 (10) | 65 (10) | 61 (11) |
| Gracilis Index ($ml/m^2$) | 59 (13) | 68 (13) | 64 (14) |
| Pectineus Index ($ml/m^2$) | 36.6 (6.6) | 33.9 (5.0) | 35.2 (6.0) |
| Pelvic Floor Index ($ml/m^2$) | 44.1 (5.0) | 42.8 (4.6) | 43.4 (4.8) |
| Piriformis Index ($ml/m^2$) | 32.4 (6.3) | 29.7 (5.0) | 31.0 (5.8) |
| Rectus Femoris Index ($ml/m^2$) | 125 (19) | 153 (25) | 140 (26) |
| Sartorius Index ($ml/m^2$) | 94 (19) | 104 (16) | 99 (18) |
| Semimembranosus Index ($ml/m^2$) | 136 (25) | 164 (28) | 151 (30) |
| Semitendinosus Index ($ml/m^2$) | 90 (17) | 110 (19) | 101 (21) |
| Short External Rotators Index ($ml/m^2$) | 33 (7) | 34 (6) | 33 (6) |
| Tensor Fasciae Latae Index ($ml/m^2$) | 47 (13) | 51 (11) | 49 (13) |
| Vastus Intermedius Index ($ml/m^2$) | 247 (38) | 315 (47) | 282 (55) |
| Vastus Lateralis Index ($ml/m^2$) | 281 (46) | 358 (53) | 321 (62) |
| Vastus Medialis Index ($ml/m^2$) | 214 (32) | 260 (37) | 238 (42) |

[1]Mean (SD)

**Table S1.** Average total volume divided by height squared of the twenty bilateral muscle groups in the hips and thighs, separated by sex. Values are reported as mean and standard deviation.

| Characteristic | Women | | Men | |
|---|---|---|---|---|
| | baseline<br>$N = 1,452$[1] | re-imaging<br>$N = 1,452$[1] | baseline<br>$N = 1,314$[1] | re-imaging<br>$N = 1,314$[1] |
| Age (years) | 62 (7) | 65 (7) | 64 (7) | 66 (7) |
| White Ethnicity | 1,269 (87%) | 1,269 (87%) | 1,171 (89%) | 1,171 (89%) |
| Weight (kg) | 68 (12) | 68 (12) | 82 (13) | 82 (13) |
| Height (m) | 1.63 (0.06) | 1.63 (0.06) | 1.75 (0.06) | 1.75 (0.06) |
| BMI ($kg/m^2$) | 25.7 (4.3) | 25.8 (4.4) | 26.7 (3.8) | 26.7 (3.8) |
| Waist Circumference (cm) | 83 (11) | 83 (11) | 93 (11) | 94 (11) |
| Hip Circumference (cm) | 101 (9) | 100 (9) | 100 (7) | 99 (7) |
| Waist-to-Hip Ratio | 0.82 (0.07) | 0.83 (0.07) | 0.93 (0.06) | 0.94 (0.07) |
| Dominant Hand Grip Strength (kg) | 24 (6) | 22 (6) | 39 (8) | 36 (9) |
| Systolic Blood Pressure (mmHg) | 135 (18) | 139 (19) | 142 (16) | 144 (17) |
| Diastolic Blood Pressure (mmHg) | 76 (10) | 78 (10) | 81 (10) | 81 (10) |
| Total MET (hours/week) | 49 (40) | 50 (41) | 52 (44) | 51 (43) |
| Sedentary Time (hours) | 4.89 (2.02) | 5.03 (2.09) | 5.47 (2.19) | 5.63 (2.21) |
| T2D | 48 (3.3%) | 20 (1.4%) | 84 (6.4%) | 30 (2.3%) |
| ASAT Volume (l) | 9.9 (4.1) | 9.9 (4.2) | 7.0 (3.3) | 7.0 (3.3) |
| VAT Volume (l) | 2.87 (1.53) | 2.94 (1.56) | 5.21 (2.33) | 5.27 (2.36) |

[1]Mean (SD); n (%)

**Table S2.** Demographics for participants in the longitudinal cohort, separated by sex. Values for continuous variables are reported as mean and standard deviation, values for discrete variables are reported as percentages. ASAT: abdominal subcutaneous adipose tissue; BMI: body mass index; MET: metabolic equivalent of task; T2D: type-2 diabetes; VAT: visceral adipose tissue.

| IDP | Women | | Men | | | | | | | |
|---|---|---|---|---|---|---|---|---|---|---|
| | baseline $N = 1,452$[1] | re-imaging $N = 1,452$[1] | baseline $N = 1,314$[1] | re-imaging $N = 1,314$[1] | Δ (Women) | Δ (Men) | %Δ (Women) | %Δ (Men) | $p$ (Women)[2] | $p$ (Men)[2] |
| Adductor Brevis (ml) | 105 (20) | 106 (20) | 163 (27) | 166 (27) | 0.7 (16.4) | 2.9 (15.0) | 2.1 (16.4) | 2.3 (9.8) | 0.0575 | < 0.0001 |
| Adductor Longus (ml) | 243 (52) | 239 (52) | 325 (57) | 321 (58) | −3.6 (22.7) | −3.7 (20.4) | −1.2 (9.4) | −1.0 (6.4) | < 0.0001 | < 0.0001 |
| Adductor Magnus (ml) | 886 (150) | 883 (149) | 1,338 (210) | 1,319 (209) | −3.6 (40.2) | −18.4 (55.4) | −0.3 (4.8) | −1.3 (4.1) | 0.0007 | < 0.0001 |
| Biceps Femoris (ml) | 457 (79) | 454 (78) | 651 (105) | 643 (104) | −3.0 (23.4) | −7.8 (49.9) | −0.5 (5.2) | −0.3 (23.4) | < 0.0001 | < 0.0001 |
| Gluteus Maximus (ml) | 1,406 (243) | 1,393 (244) | 1,909 (317) | 1,876 (315) | −12.8 (59.6) | −33.5 (83.0) | −0.9 (4.1) | −1.7 (4.2) | < 0.0001 | < 0.0001 |
| Gluteus Medius (ml) | 572 (81) | 566 (81) | 749 (99) | 741 (99) | −5.3 (19.3) | −8.0 (24.5) | −0.9 (3.4) | −1.0 (3.3) | < 0.0001 | < 0.0001 |
| Gluteus Minimus (ml) | 155 (28) | 151 (30) | 199 (32) | 196 (33) | −3.8 (7.7) | −2.5 (8.1) | −2.6 (5.5) | −1.3 (4.6) | < 0.0001 | < 0.0001 |
| Gracilis (ml) | 152 (33) | 151 (33) | 208 (40) | 206 (40) | −0.9 (9.7) | −2.6 (15.7) | −0.4 (6.7) | −1.0 (8.4) | 0.0004 | < 0.0001 |
| Pectineus (ml) | 95 (17) | 96 (17) | 102 (15) | 101 (15) | 0.9 (9.4) | −0.8 (7.8) | 1.4 (9.6) | −0.5 (7.8) | < 0.0001 | < 0.0001 |
| Pelvic Floor (ml) | 117 (15) | 118 (16) | 131 (15) | 130 (15) | 0.6 (4.8) | −0.7 (5.2) | 0.5 (4.3) | −0.4 (4.0) | < 0.0001 | < 0.0001 |
| Piriformis (ml) | 87 (17) | 86 (18) | 92 (16) | 91 (16) | −0.6 (5.0) | −1.0 (3.6) | −0.6 (7.3) | −1.1 (4.0) | 0.0005 | < 0.0001 |
| Rectus Femoris (ml) | 336 (57) | 331 (56) | 482 (84) | 472 (83) | −4.5 (15.7) | −9.8 (26.1) | −1.2 (4.8) | −1.9 (5.6) | < 0.0001 | < 0.0001 |
| Sartorius (ml) | 238 (45) | 241 (45) | 312 (51) | 313 (52) | 3.0 (13.5) | 1.9 (20.3) | 1.5 (6.3) | 0.8 (7.3) | < 0.0001 | 0.0005 |
| Semimembranosus (ml) | 362 (65) | 350 (65) | 503 (90) | 491 (90) | −11.7 (24.5) | −11.9 (48.6) | −3.1 (6.8) | 1.8 (95.6) | < 0.0001 | < 0.0001 |
| Semitendinosus (ml) | 237 (44) | 235 (45) | 340 (63) | 336 (63) | −2.6 (15.4) | −3.7 (22.1) | −1.0 (6.7) | −1.0 (7.2) | < 0.0001 | < 0.0001 |
| Short External Rotators (ml) | 88 (18) | 88 (18) | 102 (20) | 101 (20) | 0.4 (5.1) | −1.2 (6.9) | 0.6 (6.5) | −0.8 (7.3) | 0.0049 | < 0.0001 |
| Tensor Fasciae (ml) | 119 (32) | 120 (34) | 155 (35) | 153 (36) | 0.6 (7.6) | −2.1 (9.9) | 0.3 (6.2) | −1.3 (6.9) | 0.0039 | < 0.0001 |
| Vastus Intermedius (ml) | 654 (109) | 640 (108) | 975 (157) | 959 (158) | −13.5 (34.1) | −16.2 (52.8) | −2.0 (5.4) | −1.5 (6.0) | < 0.0001 | < 0.0001 |
| Vastus Lateralis (ml) | 750 (129) | 736 (128) | 1,100 (179) | 1,084 (182) | −14.2 (41.1) | −16.5 (74.3) | −1.8 (5.7) | −1.3 (7.4) | < 0.0001 | < 0.0001 |
| Vastus Medialis (ml) | 570 (95) | 559 (96) | 804 (129) | 790 (130) | −11.4 (35.2) | −14.5 (64.8) | −1.9 (6.5) | −1.0 (22.5) | < 0.0001 | < 0.0001 |

[1]Mean (SD)
[2]Wilcoxon signed-rank test

**Table S3.** Summary of total leg-muscle volume IDPs at the baseline and re-imaging visits for the longitudinal cohort, separated by sex. Values are reported as mean and standard deviation. Differences are provided in both raw units ($\Delta$ = re-imaging − baseline [in ml]) and as percentages ($\%\Delta = [\text{re-imaging} - \text{baseline}]/\text{baseline} \times 100$). The Bonferroni corrected p-value threshold for statistical significance is 0.00010417.

| IDP | Women baseline $N = 1,452$[1] | Women re-imaging $N = 1,452$[1] | Men baseline $N = 1,314$[1] | Men re-imaging $N = 1,314$[1] | Δ (Women) | Δ (Men) | %Δ (Women) | %Δ (Men) | $p$ (Women)[2] | $p$ (Men)[2] |
|---|---|---|---|---|---|---|---|---|---|---|
| Adductor Brevis (%) | 7.88 (3.04) | 8.03 (3.22) | 4.95 (1.29) | 5.04 (1.47) | 0.2 (1.7) | 0.1 (0.6) | 3.3 (19.7) | 1.6 (11.0) | 0.0004 | < 0.0001 |
| Adductor Longus (%) | 5.66 (1.90) | 5.85 (2.08) | 5.46 (2.15) | 5.64 (2.42) | 0.2 (0.7) | 0.2 (0.7) | 3.3 (11.8) | 2.8 (12.2) | < 0.0001 | < 0.0001 |
| Adductor Magnus (%) | 6.01 (2.38) | 6.23 (2.52) | 5.19 (1.82) | 5.30 (2.04) | 0.2 (0.9) | 0.1 (0.6) | 3.5 (13.0) | 1.7 (10.6) | < 0.0001 | < 0.0001 |
| Biceps Femoris (%) | 9.3 (4.2) | 9.9 (4.5) | 7.5 (3.4) | 8.0 (3.7) | 0.6 (1.4) | 0.4 (1.1) | 8.0 (15.4) | 6.2 (13.3) | < 0.0001 | < 0.0001 |
| Gluteus Maximus (%) | 18 (8) | 19 (9) | 14 (7) | 15 (8) | 0.8 (1.8) | 0.7 (1.7) | 4.9 (10.8) | 4.7 (11.5) | < 0.0001 | < 0.0001 |
| Gluteus Medius (%) | 8.7 (3.8) | 9.2 (4.1) | 7.9 (3.2) | 8.1 (3.5) | 0.4 (1.1) | 0.2 (1.0) | 4.9 (12.1) | 2.7 (10.5) | < 0.0001 | < 0.0001 |
| Gluteus Minimus (%) | 13 (7) | 14 (8) | 8.3 (4.0) | 8.6 (4.4) | 1.0 (1.7) | 0.4 (1.2) | 8.3 (13.2) | 4.2 (12.2) | < 0.0001 | < 0.0001 |
| Gracilis (%) | 18 (7) | 19 (7) | 8.2 (3.4) | 8.6 (3.7) | 0.4 (2.6) | 0.3 (1.3) | 3.2 (14.9) | 4.7 (14.6) | < 0.0001 | < 0.0001 |
| Pectineus (%) | 9.20 (2.93) | 9.51 (3.10) | 5.84 (1.45) | 5.98 (1.66) | 0.3 (1.1) | 0.1 (0.7) | 3.8 (11.6) | 2.5 (11.4) | < 0.0001 | < 0.0001 |
| Pelvic Floor (%) | 16 (6) | 17 (6) | 9.1 (3.5) | 9.5 (3.8) | 0.9 (2.1) | 0.4 (1.4) | 6.9 (14.3) | 5.2 (14.5) | < 0.0001 | < 0.0001 |
| Piriformis (%) | 17.4 (5.7) | 18.2 (5.8) | 11.2 (3.9) | 11.7 (4.2) | 0.8 (1.9) | 0.5 (1.4) | 5.1 (11.6) | 4.4 (11.5) | < 0.0001 | < 0.0001 |
| Rectus Femoris (%) | 6.95 (2.72) | 7.17 (2.91) | 5.14 (1.25) | 5.21 (1.44) | 0.2 (0.9) | 0.1 (0.5) | 3.2 (11.7) | 1.1 (8.8) | < 0.0001 | 0.0258 |
| Sartorius (%) | 19 (7) | 20 (7) | 9.9 (4.5) | 10.5 (4.9) | 1.1 (2.3) | 0.6 (1.4) | 7.1 (13.8) | 7.1 (14.4) | < 0.0001 | < 0.0001 |
| Semimembranosus (%) | 11.6 (6.0) | 12.5 (6.5) | 9.9 (5.2) | 10.6 (5.7) | 0.9 (1.7) | 0.7 (1.6) | 8.8 (15.1) | 7.1 (14.1) | < 0.0001 | < 0.0001 |
| Semitendinosus (%) | 9.3 (4.0) | 9.7 (4.2) | 7.13 (2.94) | 7.38 (3.21) | 0.4 (1.3) | 0.3 (1.0) | 4.3 (13.3) | 3.4 (11.7) | < 0.0001 | < 0.0001 |
| Short External Rotators (%) | 13 (6) | 14 (7) | 7.1 (3.3) | 7.3 (3.7) | 0.5 (1.8) | 0.2 (1.1) | 3.8 (13.1) | 2.4 (12.5) | < 0.0001 | < 0.0001 |
| Tensor Fasciae (%) | 23 (10) | 23 (10) | 14 (7) | 14 (8) | 0.8 (2.4) | 0.7 (2.0) | 4.1 (11.9) | 5.0 (13.5) | < 0.0001 | < 0.0001 |
| Vastus Intermedius (%) | 4.62 (1.91) | 4.71 (2.10) | 4.31 (1.35) | 4.32 (1.52) | 0.1 (0.7) | 0.0 (0.5) | 1.3 (13.4) | −0.3 (10.0) | 0.0399 | 0.0004 |
| Vastus Lateralis (%) | 5.35 (2.43) | 5.61 (2.70) | 4.54 (1.41) | 4.63 (1.59) | 0.3 (0.8) | 0.1 (0.6) | 4.6 (15.1) | 1.8 (12.1) | < 0.0001 | 0.0005 |
| Vastus Medialis (%) | 5.04 (2.26) | 5.29 (2.53) | 4.16 (1.45) | 4.25 (1.68) | 0.2 (0.8) | 0.1 (0.5) | 4.4 (14.4) | 1.3 (11.3) | < 0.0001 | 0.0178 |

[1] Mean (SD)
[2] Wilcoxon signed-rank test

**Table S4.** Summary of relative fat fraction IDPs at the baseline and re-imaging visits for the longitudinal cohort, separated by sex. Values are reported as mean and standard deviation. Differences are provided in both raw units (Δ = re-imaging − baseline [in ml]) and as percentages (%Δ = [re-imaging − baseline]/baseline × 100). The Bonferroni corrected p-value threshold for statistical significance is 0.00010417.

| Characteristic | Women | | Men | |
|---|---|---|---|---|
| | T2D<br>n = 628[1] | control<br>n = 628[1] | T2D<br>n = 1,088[1] | control<br>n = 1,088[1] |
| Age (years) | 66 (7) | 66 (7) | 67 (7) | 67 (7) |
| White Ethnicity | 553 (88%) | 582 (93%) | 961 (88%) | 1,012 (93%) |
| Weight (kg) | 79 (16) | 78 (16) | 89 (14) | 89 (14) |
| Height (m) | 1.62 (0.06) | 1.61 (0.06) | 1.74 (0.06) | 1.75 (0.06) |
| BMI (kg/m$^2$) | 30.0 (5.6) | 30.0 (5.6) | 29.1 (4.3) | 29.1 (4.3) |
| Waist Circumference (cm) | 94 (13) | 91 (13) | 101 (11) | 99 (11) |
| Hip Circumference (cm) | 107 (12) | 108 (12) | 103 (8) | 104 (8) |
| Waist-to-Hip Ratio | 0.88 (0.08) | 0.85 (0.07) | 0.98 (0.07) | 0.96 (0.06) |
| Dominant Hand Grip Strength (kg) | 22 (6) | 23 (6) | 36 (9) | 38 (8) |
| Systolic Blood Pressure (mmHg) | 141 (19) | 142 (19) | 143 (17) | 146 (18) |
| Diastolic Blood Pressure (mmHg) | 76 (10) | 79 (10) | 78 (10) | 82 (10) |
| Total MET (hours/week) | 43 (42) | 43 (40) | 43 (44) | 44 (40) |
| Sedentary Time (hours) | 5.93 (2.33) | 5.33 (2.07) | 6.34 (2.44) | 5.89 (2.28) |
| GLP-1 intake | 7 (1.1%) | 0 (0%) | 23 (2.1%) | 0 (0%) |
| T2D | 628 (100%) | 0 (0%) | 1,088 (100%) | 0 (0%) |
| ASAT Volume (l) | 13.4 (5.2) | 13.5 (5.3) | 8.7 (3.7) | 8.7 (3.9) |
| VAT Volume (l) | 4.60 (1.83) | 4.09 (1.78) | 7.00 (2.53) | 6.46 (2.43) |

[1]Mean (SD); n (%)

**Table S5.** Demographics for participants in the type-2 diabetes case-cohort cohort, separated by sex. Values for continuous variables are reported as mean and standard deviation, values for discrete variables are reported as percentages. ASAT: abdominal subcutaneous adipose tissue; BMI: body mass index; MET: metabolic equivalent of task; VAT: visceral adipose tissue.

| IDP | Women T2D $n = 628$[1] | Women control $n = 628$[1] | Women p-value[2] | Men T2D $n = 1,088$[1] | Men control $n = 1,088$[1] | Men p-value[2] |
|---|---|---|---|---|---|---|
| Adductor Brevis (ml) | 108 (24) | 105 (22) | 0.0082 | 166 (29) | 171 (28) | < 0.0001 |
| Adductor Longus (ml) | 255 (53) | 264 (51) | 0.0032 | 309 (54) | 329 (55) | < 0.0001 |
| Adductor Magnus (ml) | 953 (174) | 940 (173) | 0.2370 | 1,349 (216) | 1,378 (210) | 0.0007 |
| Biceps Femoris (ml) | 484 (87) | 480 (86) | 0.3931 | 653 (109) | 669 (104) | 0.0002 |
| Gluteus Maximus (ml) | 1,539 (297) | 1,518 (289) | 0.1717 | 1,897 (356) | 1,969 (337) | < 0.0001 |
| Gluteus Medius (ml) | 595 (92) | 589 (88) | 0.3979 | 749 (107) | 765 (101) | 0.0009 |
| Gluteus Minimus (ml) | 153 (32) | 150 (31) | 0.1750 | 197 (35) | 200 (35) | 0.0580 |
| Gracilis (ml) | 167 (36) | 167 (38) | 0.6755 | 205 (43) | 216 (41) | < 0.0001 |
| Pectineus (ml) | 103 (19) | 105 (17) | 0.2645 | 105 (16) | 106 (16) | 0.2112 |
| Pelvic Floor (ml) | 117 (16) | 120 (15) | 0.0013 | 130 (16) | 133 (16) | 0.0001 |
| Piriformis | 87 (17) | 89 (17) | 0.0165 | 90 (17) | 92 (16) | 0.0004 |
| Rectus Femoris (ml) | 322 (55) | 334 (56) | 0.0001 | 442 (77) | 475 (80) | < 0.0001 |
| Sartorious | 267 (51) | 268 (51) | 0.8227 | 320 (52) | 331 (51) | < 0.0001 |
| Semimembranosus (ml) | 378 (78) | 371 (77) | 0.1147 | 505 (95) | 514 (90) | 0.0023 |
| Semitendinosus (ml) | 248 (47) | 250 (48) | 0.1330 | 331 (62) | 348 (61) | < 0.0001 |
| Short External Rotators (ml) | 88 (19) | 87 (18) | 0.1404 | 102 (21) | 104 (21) | 0.0297 |
| Tensor Fasciae (ml) | 131 (37) | 135 (38) | 0.0237 | 152 (35) | 160 (36) | < 0.0001 |
| Vastus Intermedius (ml) | 667 (122) | 669 (118) | 0.9011 | 941 (162) | 980 (155) | < 0.0001 |
| Vastus Lateralis (ml) | 759 (141) | 773 (137) | 0.1453 | 1,066 (181) | 1,118 (173) | < 0.0001 |
| Vastus Medialis (ml) | 571 (100) | 576 (95) | 0.4467 | 776 (131) | 804 (124) | < 0.0001 |

[1]Mean (SD)
[2]Wilcoxon rank sum test

**Table S6.** Summary of total leg-muscle volume for the type-2 diabetes case-control cohort, separated by sex. Values are reported as mean and standard deviation. The Bonferroni corrected p-value threshold for statistical significance is 0.00010417.

| IDP | Women | | | Men | | |
|---|---|---|---|---|---|---|
| | T2D $n = 628$[1] | control $n = 628$[1] | p-value[2] | T2D $n = 1,088$[1] | control $n = 1,088$[1] | p-value[2] |
| Adductor Brevis (%) | 10.0 (4.10) | 9.8 (3.68) | 0.9601 | 6.0 (2.00) | 5.5 (1.62) | < 0.0001 |
| Adductor Longus (%) | 7.6 (3.03) | 6.9 (2.45) | 0.0002 | 7.1 (3.38) | 6.4 (2.84) | < 0.0001 |
| Adductor Magnus (%) | 7.9 (4.02) | 7.6 (3.49) | 0.2943 | 6.5 (3.17) | 6.1 (2.56) | 0.0050 |
| Biceps Femoris (%) | 13.6 (6.79) | 12.9 (6.23) | 0.1524 | 10.2 (5.42) | 9.3 (4.53) | 0.0010 |
| Gluteus Maximus (%) | 26.8 (11.51) | 24.1 (10.09) | < 0.0001 | 20.9 (10.01) | 18.1 (8.84) | < 0.0001 |
| Gluteus Medius (%) | 13.3 (6.02) | 12.0 (5.63) | < 0.0001 | 11.0 (5.56) | 9.5 (4.09) | < 0.0001 |
| Gluteus Minimus (%) | 17.1 (9.11) | 17.6 (9.22) | 0.3264 | 10.6 (5.84) | 9.7 (4.92) | 0.0002 |
| Gracilis (%) | 20.2 (7.80) | 20.5 (7.39) | 0.5473 | 10.5 (4.79) | 9.4 (4.21) | < 0.0001 |
| Pectineus (%) | 10.9 (3.93) | 10.3 (3.13) | 0.0824 | 7.0 (2.14) | 6.5 (1.85) | < 0.0001 |
| Pelvic Floor (%) | 20.7 (6.95) | 20.1 (6.68) | 0.1325 | 11.7 (4.72) | 10.6 (4.18) | < 0.0001 |
| Piriformis (%) | 20.9 (6.29) | 20.5 (5.91) | 0.3114 | 14.0 (5.27) | 12.7 (4.43) | < 0.0001 |
| Rectus Femoris (%) | 9.7 (4.32) | 9.1 (4.18) | 0.0208 | 6.5 (2.43) | 5.8 (1.86) | < 0.0001 |
| Sartorious (%) | 23.0 (8.35) | 22.7 (7.53) | 0.6009 | 13.2 (6.20) | 12.1 (5.66) | < 0.0001 |
| Semimembranosus (%) | 17.8 (9.40) | 16.7 (9.00) | 0.0228 | 14.0 (7.79) | 12.5 (6.57) | < 0.0001 |
| Semitendinosus (%) | 13.0 (6.16) | 12.1 (5.56) | 0.0411 | 9.3 (4.56) | 8.6 (4.13) | < 0.0001 |
| Short External Rotators (%) | 16.7 (8.14) | 17.0 (7.68) | 0.1553 | 9.3 (5.01) | 8.6 (4.46) | 0.0003 |
| Tensor Fasciae (%) | 30.5 (11.72) | 28.2 (11.06) | 0.0008 | 19.1 (9.47) | 17.4 (8.47) | < 0.0001 |
| Vastus Intermedius (%) | 6.6 (3.30) | 6.2 (3.00) | 0.0027 | 5.5 (2.70) | 5.1 (2.02) | < 0.0001 |
| Vastus Lateralis (%) | 7.9 (4.23) | 7.5 (3.87) | 0.0593 | 5.9 (2.86) | 5.3 (2.16) | < 0.0001 |
| Vastus Medialis (%) | 7.6 (3.96) | 7.0 (3.55) | 0.0020 | 5.5 (2.80) | 4.9 (2.15) | < 0.0001 |

[1] Mean (SD)
[2] Wilcoxon rank sum test

**Table S7.** Median relative fat fraction (rFF) of the twenty bilateral muscle groups in the hips and thighs for the type-2 diabetes case-control cohort, separated by sex. Values are reported as mean and standard deviation. The Bonferroni corrected p-value threshold for statistical significance is 0.00010417.

| Characteristic | Women | | Men | |
|---|---|---|---|---|
| | control $n = 11$[1] | GLP-1RA $n = 11$[1] | control $n = 27$[1] | GLP-1RA $n = 27$[1] |
| Age (years) | 69 (7) | 69 (7) | 69 (5) | 69 (5) |
| White Ethnicity | 9 (82%) | 10 (91%) | 25 (93%) | 21 (78%) |
| Weight (kg) | 84 (12) | 85 (12) | 94 (12) | 95 (13) |
| Height (m) | 1.61 (0.06) | 1.62 (0.05) | 1.74 (0.06) | 1.75 (0.05) |
| BMI (kg/m$^2$) | 32.4 (3.4) | 32.5 (3.6) | 31.1 (3.5) | 31.3 (3.6) |
| Waist Circumference (cm) | 101 (10) | 103 (10) | 108 (11) | 107 (11) |
| Hip Circumference (cm) | 111 (7) | 112 (9) | 105 (7) | 104 (7) |
| Waist-to-Hip Ratio | 0.91 (0.05) | 0.92 (0.06) | 1.03 (0.07) | 1.03 (0.08) |
| Dominant Hand Grip Strength (kg) | 20.5 (3.3) | 18.9 (4.5) | 34 (9) | 35 (5) |
| Systolic Blood Pressure (mmHg) | 149 (21) | 151 (22) | 147 (21) | 140 (16) |
| Diastolic Blood Pressure (mmHg) | 76 (9) | 81 (9) | 81 (10) | 76 (6) |
| Townsend Deprivation Index | −1.90 (2.73) | −1.80 (2.71) | −1.91 (2.40) | −1.81 (2.30) |
| Total MET (hours/week) | 50 (62) | 93 (93) | 49 (49) | 39 (41) |
| Sedentary Time (hours) | 5.59 (2.83) | 5.91 (1.80) | 6.15 (1.48) | 6.63 (1.68) |
| ASAT Volume (l) | 14.8 (5.3) | 15.5 (3.7) | 10.2 (3.1) | 10.8 (3.7) |
| VAT Volume (l) | 5.97 (2.21) | 5.73 (1.28) | 8.29 (2.62) | 8.59 (2.27) |
| Duration of GLP-1RA Treatment (months) | 0.00 (–) | 6.27 (2.76) | 0.00 (–) | 6.36 (2.67) |

[1] Mean (SD); n (%)

**Table S8.** Demographics for participants in the GLP-1RA case-control cohort, separated by sex. Values for continuous variables are reported as mean and standard deviation, values for discrete variables are reported as percentages. ASAT: abdominal subcutaneous adipose tissue; BMI: body mass index; GLP-1RA - glucagon-like peptide-1 receptor agonist; MET: metabolic equivalent of task; VAT: visceral adipose tissue.

| IDP | Women | | | Men | | |
|---|---|---|---|---|---|---|
| | control $n = 11$[1] | GLP-1RA $n = 11$[1] | p-value[2] | control $n = 27$[1] | GLP-1RA $n = 27$[1] | p-value[2] |
| Adductor Brevis (ml) | 100 (12) | 112 (18) | 0.0652 | 170 (24) | 159 (35) | 0.2706 |
| Adductor Longus (ml) | 256 (46) | 279 (53) | 0.2426 | 321 (53) | 289 (64) | 0.0460 |
| Adductor Magnus (ml) | 964 (233) | 992 (202) | 0.6063 | 1,389 (230) | 1,357 (233) | 0.5710 |
| Biceps Femoris (ml) | 483 (83) | 501 (86) | 0.4779 | 690 (107) | 643 (107) | 0.0638 |
| Gluteus Maximus (ml) | 1,539 (290) | 1,512 (147) | 0.5619 | 1,996 (324) | 1,912 (372) | 0.4094 |
| Gluteus Medius (ml) | 551 (91) | 623 (39) | 0.0281 | 798 (103) | 735 (123) | 0.0372 |
| Gluteus Minimus (ml) | 141 (36) | 149 (26) | 0.3000 | 200 (40) | 193 (39) | 0.7058 |
| Gracilis (ml) | 183 (47) | 189 (48) | 0.5190 | 220 (45) | 210 (44) | 0.2706 |
| Pectineus (ml) | 109 (18) | 107 (18) | 0.9487 | 111 (15) | 104 (15) | 0.2464 |
| Pelvic Floor (ml) | 113 (17) | 123 (12) | 0.1164 | 133 (13) | 129 (24) | 0.1377 |
| Piriformis (ml) | 81 (15) | 79 (21) | 0.9487 | 92 (19) | 81 (20) | 0.1287 |
| Rectus Femoris (ml) | 312 (33) | 318 (44) | 0.4385 | 461 (70) | 388 (82) | 0.0026 |
| Sartorious (ml) | 292 (39) | 284 (51) | 0.6994 | 341 (45) | 318 (54) | 0.0868 |
| Semimembranosus (ml) | 357 (75) | 390 (100) | 0.2169 | 519 (101) | 498 (111) | 0.4706 |
| Semitendinosus (ml) | 262 (46) | 279 (55) | 0.6522 | 345 (58) | 325 (64) | 0.1902 |
| Short External Rotators (ml) | 88 (12) | 94 (6) | 0.3653 | 108 (18) | 100 (20) | 0.1572 |
| Tensor Fasciae (ml) | 131 (32) | 131 (42) | 0.9487 | 169 (42) | 150 (33) | 0.1472 |
| Vastus Intermedius (ml) | 643 (141) | 658 (98) | 0.3653 | 921 (138) | 946 (166) | 0.2939 |
| Vastus Lateralis (ml) | 742 (156) | 717 (127) | 0.2703 | 1,080 (153) | 1,046 (205) | 0.6186 |
| Vastus Medialis (ml) | 551 (114) | 552 (75) | 0.4385 | 775 (116) | 763 (131) | 0.9931 |

[1]Mean (SD)
[2]Wilcoxon rank sum test

**Table S9.** Summary of total leg-muscle volume for the GLP-1RA case-control cohort, separated by sex. Values are reported as mean and standard deviation. The Bonferroni corrected p-value threshold for statistical significance is 0.00010417.

| IDP | Women | | | Men | | |
|---|---|---|---|---|---|---|
| | control<br>$n = 11$[1] | GLP-1RA<br>$n = 11$[1] | p-value[2] | control<br>$n = 27$[1] | GLP-1RA<br>$n = 27$[1] | p-value[2] |
| Adductor Brevis (%) | 12.0 (4.72) | 8.0 (1.78) | 0.0192 | 5.5 (1.28) | 6.6 (2.94) | 0.2486 |
| Adductor Longus (%) | 8.9 (4.35) | 5.9 (1.24) | 0.0128 | 7.2 (2.48) | 8.3 (3.81) | 0.5029 |
| Adductor Magnus (%) | 9.5 (5.47) | 6.2 (1.55) | 0.0879 | 6.1 (2.19) | 7.9 (4.91) | 0.2193 |
| Biceps Femoris (%) | 18.8 (9.26) | 10.7 (3.35) | 0.0158 | 9.9 (4.53) | 13.5 (7.64) | 0.0691 |
| Gluteus Maximus (%) | 32.3 (12.46) | 22.9 (5.61) | 0.0400 | 20.5 (8.30) | 27.3 (12.38) | 0.0388 |
| Gluteus Medius (%) | 19.4 (10.52) | 10.7 (2.39) | 0.0473 | 10.6 (4.18) | 13.2 (5.62) | 0.0852 |
| Gluteus Minimus (%) | 23.7 (10.98) | 15.5 (5.20) | 0.0759 | 10.7 (5.57) | 12.5 (6.04) | 0.1902 |
| Gracilis (%) | 24.1 (12.93) | 15.7 (6.00) | 0.1932 | 9.8 (4.34) | 13.8 (8.20) | 0.1331 |
| Pectineus (%) | 12.7 (5.46) | 9.7 (2.30) | 0.2426 | 6.6 (1.81) | 7.8 (2.34) | 0.0693 |
| Pelvic Floor (%) | 25.3 (8.18) | 18.8 (5.19) | 0.0557 | 11.6 (3.69) | 13.1 (5.88) | 0.4094 |
| Piriformis (%) | 24.7 (9.56) | 20.5 (6.39) | 0.3000 | 13.7 (4.76) | 17.3 (6.96) | 0.0441 |
| Rectus Femoris (%) | 12.7 (5.46) | 8.1 (2.91) | 0.0104 | 5.6 (1.53) | 7.6 (3.18) | 0.0035 |
| Sartorious (%) | 27.2 (9.97) | 18.6 (7.77) | 0.0400 | 12.3 (4.25) | 15.7 (7.43) | 0.1287 |
| Semimembranosus (%) | 25.3 (11.43) | 15.1 (5.59) | 0.0128 | 15.0 (7.36) | 18.7 (9.29) | 0.1472 |
| Semitendinosus (%) | 16.0 (9.20) | 9.3 (3.68) | 0.0473 | 8.6 (3.58) | 11.8 (7.45) | 0.0970 |
| Short External Rotators (%) | 20.8 (10.60) | 12.6 (4.79) | 0.0473 | 8.6 (4.01) | 10.9 (5.88) | 0.0868 |
| Tensor Fasciae (%) | 35.4 (10.97) | 25.4 (9.60) | 0.0557 | 18.1 (8.85) | 23.8 (10.84) | 0.0260 |
| Vastus Intermedius (%) | 8.6 (4.13) | 5.7 (1.40) | 0.0759 | 5.4 (2.24) | 5.9 (2.37) | 0.4600 |
| Vastus Lateralis (%) | 9.8 (5.54) | 7.1 (2.12) | 0.2703 | 5.3 (1.77) | 6.7 (3.10) | 0.0720 |
| Vastus Medialis (%) | 9.8 (4.84) | 6.8 (1.98) | 0.2426 | 5.2 (1.87) | 6.3 (3.16) | 0.2213 |

[1] Mean (SD)
[2] Wilcoxon rank sum test

**Table S10.** Median relative fat fraction of the twenty bilateral muscle groups in the hips and thighs for the GLP-1RA case-control cohort, separated by sex. Values are reported as mean and standard deviation. The Bonferroni corrected p-value threshold for statistical significance is 0.00010417.

| Sex | IDP | Intercept | Slope | p-value | $R^2$ |
|---|---|---|---|---|---|
| Female | Adductor Brevis | 23.51 | 0.62 | 0.017 | 0.406 |
| | Adductor Longus | 18.59 | 0.85 | 0.025 | 0.806 |
| | Adductor Magnus | 56.39 | 0.88 | 0.040 | 0.718 |
| | Biceps Femoris | 19.95 | 0.88 | 0.033 | 0.783 |
| | Gluteus Maximus | 31.52 | 0.98 | 0.171 | 0.883 |
| | Gluteus Medius | 36.14 | 0.85 | 0.028 | 0.773 |
| | Gluteus Minimus | 14.36 | 0.77 | 0.024 | 0.591 |
| | Gracilis | 12.16 | 0.85 | 0.033 | 0.707 |
| | Pectineus | 6.66 | 0.92 | 0.059 | 0.722 |
| | Pelvic Floor | 12.45 | 0.86 | 0.044 | 0.619 |
| | Piriformis | 8.07 | 0.79 | 0.028 | 0.586 |
| | Rectus Femoris | 17.79 | 0.91 | 0.048 | 0.792 |
| | Sartorious | 9.71 | 0.92 | 0.041 | 0.844 |
| | Semimembranosus | 14.06 | 0.93 | 0.063 | 0.786 |
| | Semitendinosus | 18.57 | 0.89 | 0.045 | 0.706 |
| | Short External Rotators | 8.25 | 0.81 | 0.029 | 0.621 |
| | Tensor Fasciae Latae | 10.53 | 0.82 | 0.027 | 0.683 |
| | Vastus Intermedius | 32.38 | 0.91 | 0.044 | 0.814 |
| | Vastus Lateralis | 28.01 | 0.88 | 0.033 | 0.792 |
| | Vastus Medialis | 38.64 | 0.89 | 0.041 | 0.747 |
| Male | Adductor Brevis | 17.50 | 0.78 | 0.021 | 0.675 |
| | Adductor Longus | 26.06 | 0.86 | 0.032 | 0.724 |
| | Adductor Magnus | 110.17 | 0.86 | 0.036 | 0.671 |
| | Biceps Femoris | 46.85 | 0.85 | 0.030 | 0.727 |
| | Gluteus Maximus | 71.84 | 0.96 | 0.090 | 0.819 |
| | Gluteus Medius | 47.25 | 0.85 | 0.027 | 0.773 |
| | Gluteus Minimus | 26.31 | 0.71 | 0.019 | 0.555 |
| | Gracilis | 24.11 | 0.78 | 0.025 | 0.587 |
| | Pectineus | 10.74 | 0.84 | 0.033 | 0.646 |
| | Pelvic Floor | 17.69 | 0.80 | 0.028 | 0.617 |
| | Piriformis | 12.57 | 0.70 | 0.019 | 0.520 |
| | Rectus Femoris | 39.13 | 0.84 | 0.033 | 0.652 |
| | Sartorious | 25.26 | 0.83 | 0.024 | 0.763 |
| | Semimembranosus | 37.71 | 0.88 | 0.038 | 0.719 |
| | Semitendinosus | 29.57 | 0.87 | 0.040 | 0.688 |
| | Short External Rotators | 14.14 | 0.77 | 0.024 | 0.593 |
| | Tensor Fasciae Latae | 14.46 | 0.81 | 0.029 | 0.637 |
| | Vastus Intermedius | 67.92 | 0.87 | 0.032 | 0.752 |
| | Vastus Lateralis | 96.44 | 0.80 | 0.024 | 0.671 |
| | Vastus Medialis | 70.56 | 0.84 | 0.029 | 0.723 |

**Table S11.** Regression coefficients and model statistics for the linear models (right volume = intercept + slope $*$ left volume) in Figures S2 and S3. The p-value is for the hypothesis test $H_0$ : slope = 1 vs $H_1$ : slope $\neq$ 1. IDP: image-derived phenotype.

| IDP | BMI Group | Sex | Intercept | Slope | p-value | $R^2$ |
|---|---|---|---|---|---|---|
| | BMI (10,20] | Women | 3.41 | −0.05 | 0.164 | 0.027 |
| | | Men | 6.92 | −0.11 | 0.071 | 0.189 |
| | BMI (20,25] | Women | 1.56 | −0.02 | 0.135 | 0.003 |
| | | Men | 1.11 | −0.01 | 0.508 | 0.001 |
| Adductor Brevis | BMI (25,30] | Women | 0.54 | −0.01 | 0.722 | 0.000 |
| | | Men | 3.22 | −0.04 | 0.001 | 0.019 |
| | BMI (30,35] | Women | −0.99 | 0.02 | 0.602 | 0.002 |
| | | Men | 2.62 | −0.04 | 0.145 | 0.011 |
| | BMI (35,Inf] | Women | 2.17 | −0.03 | 0.646 | 0.005 |
| | | Men | −0.94 | 0.02 | 0.729 | 0.004 |
| | BMI (10,20] | Women | 8.68 | −0.15 | 0.003 | 0.119 |
| | | Men | −2.60 | 0.02 | 0.830 | 0.003 |
| | BMI (20,25] | Women | 1.08 | −0.02 | 0.169 | 0.003 |
| | | Men | 2.22 | −0.04 | 0.049 | 0.009 |
| Adductor Longus | BMI (25,30] | Women | 0.74 | −0.02 | 0.439 | 0.001 |
| | | Men | 1.45 | −0.03 | 0.070 | 0.006 |
| | BMI (30,35] | Women | −1.83 | 0.02 | 0.648 | 0.001 |
| | | Men | 3.25 | −0.06 | 0.062 | 0.018 |
| | BMI (35,Inf] | Women | 19.57 | −0.31 | 0.011 | 0.129 |
| | | Men | 1.95 | −0.06 | 0.466 | 0.016 |
| | BMI (10,20] | Women | 7.57 | −0.11 | 0.239 | 0.019 |
| | | Men | −8.32 | 0.13 | 0.495 | 0.030 |
| | BMI (20,25] | Women | 1.33 | -0.02 | 0.613 | 0.000 |
| | | Men | 4.95 | −0.10 | 0.025 | 0.011 |
| Adductor Magnus | BMI (25,30] | Women | 4.73 | −0.08 | 0.056 | 0.007 |
| | | Men | 5.20 | −0.12 | 0.007 | 0.012 |
| | BMI (30,35] | Women | 8.51 | −0.15 | 0.074 | 0.020 |
| | | Men | 7.04 | −0.15 | 0.079 | 0.016 |
| | BMI (35,Inf] | Women | −5.76 | 0.04 | 0.833 | 0.001 |
| | | Men | 13.70 | −0.33 | 0.360 | 0.025 |
| | BMI (10,20] | Women | 4.76 | −0.08 | 0.133 | 0.031 |
| | | Men | 0.70 | −0.03 | 0.792 | 0.004 |
| | BMI (20,25] | Women | 0.01 | 0.00 | 0.906 | 0.000 |
| | | Men | 1.94 | −0.04 | 0.180 | 0.004 |
| Biceps Femoris | BMI (25,30] | Women | −3.80 | 0.05 | 0.028 | 0.009 |
| | | Men | 3.16 | −0.07 | 0.018 | 0.010 |
| | BMI (30,35] | Women | −1.90 | 0.01 | 0.783 | 0.000 |
| | | Men | 7.23 | −0.14 | 0.027 | 0.025 |
| | BMI (35,Inf] | Women | −5.93 | 0.09 | 0.442 | 0.013 |
| | | Men | 6.33 | −0.14 | 0.534 | 0.011 |
| | BMI (10,20] | Women | 20.51 | −0.35 | 0.011 | 0.087 |
| | | Men | −10.84 | 0.11 | 0.656 | 0.013 |
| | BMI (20,25] | Women | 4.33 | −0.08 | 0.077 | 0.005 |
| | | Men | 9.72 | −0.20 | 0.004 | 0.018 |
| Gluteus Maximus | BMI (25,30] | Women | 1.27 | −0.04 | 0.513 | 0.001 |
| | | Men | 7.04 | −0.17 | 0.006 | 0.013 |
| | BMI (30,35] | Women | 13.96 | −0.25 | 0.090 | 0.018 |
| | | Men | 8.78 | −0.21 | 0.197 | 0.009 |
| | BMI (35,Inf] | Women | 6.62 | −0.13 | 0.690 | 0.003 |
| | | Men | −11.67 | 0.00 | 0.996 | 0.000 |
| | BMI (10,20] | Women | 5.21 | −0.09 | 0.059 | 0.049 |
| | | Men | −0.55 | −0.01 | 0.951 | 0.000 |
| | BMI (20,25] | Women | 2.13 | −0.04 | 0.030 | 0.007 |
| Gluteus Medius | | | | | | |

| | | | | | | |
|---|---|---|---|---|---|---|
| | | Men | 1.81 | −0.04 | 0.095 | 0.006 |
| | BMI (25,30] | Women | 2.59 | −0.05 | 0.017 | 0.011 |
| | | Men | 3.24 | −0.07 | 0.002 | 0.016 |
| | BMI (30,35] | Women | 2.65 | −0.06 | 0.197 | 0.011 |
| | | Men | 6.36 | −0.12 | 0.006 | 0.038 |
| | BMI (35,Inf] | Women | 7.13 | −0.14 | 0.128 | 0.049 |
| | | Men | 0.24 | −0.05 | 0.664 | 0.006 |
| Gluteus Minimus | BMI (10,20] | Women | 1.81 | −0.04 | 0.033 | 0.061 |
| | | Men | −0.83 | 0.01 | 0.765 | 0.006 |
| | BMI (20,25] | Women | 1.17 | −0.03 | 0.000 | 0.022 |
| | | Men | 1.87 | −0.03 | 0.000 | 0.042 |
| | BMI (25,30] | Women | 0.73 | −0.02 | 0.010 | 0.013 |
| | | Men | 1.92 | −0.04 | 0.000 | 0.044 |
| | BMI (30,35] | Women | −0.10 | −0.01 | 0.538 | 0.002 |
| | | Men | 0.72 | −0.02 | 0.211 | 0.008 |
| | BMI (35,Inf] | Women | 3.42 | −0.06 | 0.093 | 0.059 |
| | | Men | 3.06 | −0.06 | 0.161 | 0.057 |
| Gracilis | BMI (10,20] | Women | 2.14 | −0.03 | 0.121 | 0.033 |
| | | Men | −3.72 | 0.07 | 0.099 | 0.161 |
| | BMI (20,25] | Women | −0.81 | 0.01 | 0.117 | 0.004 |
| | | Men | −0.08 | 0.00 | 0.988 | 0.000 |
| | BMI (25,30] | Women | −0.64 | 0.01 | 0.389 | 0.001 |
| | | Men | 0.65 | −0.02 | 0.166 | 0.003 |
| | BMI (30,35] | Women | −2.13 | 0.03 | 0.176 | 0.012 |
| | | Men | 0.32 | −0.01 | 0.556 | 0.002 |
| | BMI (35,Inf] | Women | 2.25 | −0.03 | 0.514 | 0.009 |
| | | Men | −2.02 | 0.00 | 0.995 | 0.000 |
| Pectineus | BMI (10,20] | Women | 2.79 | −0.04 | 0.017 | 0.076 |
| | | Men | 0.13 | 0.00 | 0.938 | 0.000 |
| | BMI (20,25] | Women | 0.61 | −0.01 | 0.368 | 0.001 |
| | | Men | 0.32 | 0.00 | 0.553 | 0.001 |
| | BMI (25,30] | Women | 0.25 | 0.00 | 0.930 | 0.000 |
| | | Men | −0.39 | 0.00 | 0.528 | 0.001 |
| | BMI (30,35] | Women | 1.20 | −0.01 | 0.475 | 0.003 |
| | | Men | 1.07 | −0.02 | 0.160 | 0.010 |
| | BMI (35,Inf] | Women | 2.76 | −0.04 | 0.285 | 0.024 |
| | | Men | 0.17 | −0.01 | 0.812 | 0.002 |
| Pelvic Floor | BMI (10,20] | Women | 1.31 | −0.02 | 0.211 | 0.022 |
| | | Men | −0.22 | 0.00 | 0.836 | 0.003 |
| | BMI (20,25] | Women | 0.52 | −0.01 | 0.192 | 0.003 |
| | | Men | 0.16 | 0.00 | 0.584 | 0.001 |
| | BMI (25,30] | Women | 0.28 | 0.00 | 0.787 | 0.000 |
| | | Men | −0.12 | 0.00 | 0.878 | 0.000 |
| | BMI (30,35] | Women | 0.79 | −0.01 | 0.384 | 0.005 |
| | | Men | 0.05 | 0.00 | 0.794 | 0.000 |
| | BMI (35,Inf] | Women | 0.10 | 0.00 | 0.872 | 0.001 |
| | | Men | 0.16 | 0.00 | 0.987 | 0.000 |
| Piriformis | BMI (10,20] | Women | 2.45 | −0.04 | 0.005 | 0.103 |
| | | Men | −1.25 | 0.02 | 0.392 | 0.046 |
| | BMI (20,25] | Women | 0.66 | −0.01 | 0.013 | 0.010 |
| | | Men | 0.29 | −0.01 | 0.081 | 0.007 |
| | BMI (25,30] | Women | 0.38 | −0.01 | 0.210 | 0.003 |
| | | Men | 0.13 | 0.00 | 0.219 | 0.003 |
| | BMI (30,35] | Women | 0.41 | −0.01 | 0.536 | 0.002 |
| | | Men | −0.23 | 0.00 | 0.714 | 0.001 |

| | | | | | | |
|---|---|---|---|---|---|---|
| | BMI (35,Inf] | Women | −0.32 | 0.00 | 0.760 | 0.002 |
| | | Men | −0.39 | 0.01 | 0.727 | 0.004 |
| Rectus Femoris | BMI (10,20] | Women | 9.73 | −0.16 | 0.001 | 0.136 |
| | | Men | 4.99 | −0.11 | 0.546 | 0.023 |
| | BMI (20,25] | Women | −0.68 | 0.00 | 0.791 | 0.000 |
| | | Men | 0.89 | −0.03 | 0.287 | 0.003 |
| | BMI (25,30] | Women | −3.18 | 0.04 | 0.009 | 0.013 |
| | | Men | −1.29 | 0.00 | 0.846 | 0.000 |
| | BMI (30,35] | Women | −3.05 | 0.03 | 0.308 | 0.007 |
| | | Men | −0.42 | −0.02 | 0.600 | 0.001 |
| | BMI (35,Inf] | Women | −5.69 | 0.08 | 0.355 | 0.018 |
| | | Men | −1.28 | −0.01 | 0.932 | 0.000 |
| Sartorious | BMI (10,20] | Women | 3.39 | −0.04 | 0.257 | 0.018 |
| | | Men | 4.83 | −0.07 | 0.207 | 0.097 |
| | BMI (20,25] | Women | 0.39 | 0.01 | 0.550 | 0.001 |
| | | Men | −0.51 | 0.02 | 0.298 | 0.002 |
| | BMI (25,30] | Women | −1.41 | 0.03 | 0.021 | 0.010 |
| | | Men | 2.07 | −0.03 | 0.079 | 0.005 |
| | BMI (30,35] | Women | 0.28 | 0.00 | 0.945 | 0.000 |
| | | Men | 3.63 | −0.05 | 0.056 | 0.019 |
| | BMI (35,Inf] | Women | 1.40 | −0.02 | 0.828 | 0.001 |
| | | Men | 5.93 | −0.11 | 0.168 | 0.055 |
| Semimembranosus | BMI (10,20] | Women | 4.24 | −0.10 | 0.076 | 0.043 |
| | | Men | 6.65 | −0.16 | 0.194 | 0.103 |
| | BMI (20,25] | Women | 0.26 | −0.03 | 0.215 | 0.002 |
| | | Men | 2.35 | −0.06 | 0.022 | 0.012 |
| | BMI (25,30] | Women | -0.65 | −0.02 | 0.466 | 0.001 |
| | | Men | 4.01 | −0.09 | 0.003 | 0.015 |
| | BMI (30,35] | Women | −3.35 | 0.01 | 0.880 | 0.000 |
| | | Men | 10.00 | −0.18 | 0.003 | 0.045 |
| | BMI (35,Inf] | Women | 7.34 | −0.14 | 0.171 | 0.039 |
| | | Men | 5.67 | −0.16 | 0.529 | 0.012 |
| Semitendinosus | BMI (10,20] | Women | 1.90 | −0.04 | 0.386 | 0.010 |
| | | Men | −7.01 | 0.09 | 0.336 | 0.058 |
| | BMI (20,25] | Women | −0.77 | 0.01 | 0.415 | 0.001 |
| | | Men | −0.03 | −0.01 | 0.726 | 0.000 |
| | BMI (25,30] | Women | −3.14 | 0.04 | 0.004 | 0.016 |
| | | Men | 0.59 | −0.02 | 0.329 | 0.002 |
| | BMI (30,35] | Women | −2.99 | 0.04 | 0.276 | 0.008 |
| | | Men | 1.05 | −0.02 | 0.386 | 0.004 |
| | BMI (35,Inf] | Women | −3.48 | 0.05 | 0.517 | 0.009 |
| | | Men | −2.38 | 0.02 | 0.922 | 0.000 |
| Short External Rotators | BMI (10,20] | Women | 1.95 | −0.03 | 0.100 | 0.037 |
| | | Men | 0.71 | −0.02 | 0.630 | 0.015 |
| | BMI (20,25] | Women | 0.47 | 0.00 | 0.368 | 0.001 |
| | | Men | 0.92 | −0.01 | 0.033 | 0.010 |
| | BMI (25,30] | Women | 0.29 | 0.00 | 0.463 | 0.001 |
| | | Men | 0.06 | 0.00 | 0.540 | 0.001 |
| | BMI (30,35] | Women | 0.00 | 0.00 | 0.950 | 0.000 |
| | | Men | 0.30 | −0.01 | 0.433 | 0.003 |
| | BMI (35,Inf] | Women | −2.03 | 0.03 | 0.196 | 0.035 |
| | | Men | -0.10 | 0.00 | 0.980 | 0.000 |
| Tensor Fasciae Latae | BMI (10,20] | Women | 2.85 | −0.05 | 0.001 | 0.132 |
| | | Men | 4.25 | −0.08 | 0.185 | 0.107 |
| | BMI (20,25] | Women | 0.06 | 0.00 | 0.789 | 0.000 |

| | | | | | | |
|---|---|---|---|---|---|---|
| | | Men | 0.56 | −0.01 | 0.258 | 0.003 |
| | BMI (25,30] | Women | −0.41 | 0.01 | 0.254 | 0.002 |
| | | Men | −0.67 | 0.00 | 0.593 | 0.000 |
| | BMI (30,35] | Women | −0.42 | 0.01 | 0.526 | 0.003 |
| | | Men | 0.16 | −0.01 | 0.735 | 0.001 |
| | BMI (35,Inf] | Women | −2.61 | 0.05 | 0.384 | 0.016 |
| | | Men | −3.37 | 0.05 | 0.332 | 0.028 |
| Vastus Intermedius | BMI (10,20] | Women | 6.30 | −0.11 | 0.179 | 0.025 |
| | | Men | −0.81 | −0.01 | 0.925 | 0.001 |
| | BMI (20,25] | Women | 1.52 | −0.05 | 0.110 | 0.004 |
| | | Men | 5.71 | −0.11 | 0.005 | 0.017 |
| | BMI (25,30] | Women | −1.79 | −0.01 | 0.871 | 0.000 |
| | | Men | 3.82 | −0.09 | 0.010 | 0.012 |
| | BMI (30,35] | Women | 1.78 | −0.07 | 0.261 | 0.008 |
| | | Men | 2.85 | −0.09 | 0.302 | 0.006 |
| | BMI (35,Inf] | Women | −9.20 | 0.08 | 0.612 | 0.006 |
| | | Men | −23.27 | 0.27 | 0.315 | 0.030 |
| Vastus Lateralis | BMI (10,20] | Women | 7.07 | −0.12 | 0.228 | 0.020 |
| | | Men | −22.25 | 0.32 | 0.146 | 0.127 |
| | BMI (20,25] | Women | 4.81 | −0.10 | 0.004 | 0.013 |
| | | Men | 8.17 | −0.15 | 0.011 | 0.014 |
| | BMI (25,30] | Women | −3.67 | 0.02 | 0.656 | 0.000 |
| | | Men | 4.80 | −0.11 | 0.038 | 0.007 |
| | BMI (30,35] | Women | 3.05 | −0.10 | 0.214 | 0.010 |
| | | Men | 11.48 | −0.22 | 0.099 | 0.014 |
| | BMI (35,Inf] | Women | −3.77 | 0.00 | 0.994 | 0.000 |
| | | Men | −9.80 | 0.05 | 0.912 | 0.000 |
| Vastus Medialis | BMI (10,20] | Women | 9.54 | −0.19 | 0.040 | 0.057 |
| | | Men | 1.71 | −0.06 | 0.690 | 0.010 |
| | BMI (20,25] | Women | −2.64 | 0.02 | 0.526 | 0.001 |
| | | Men | −0.06 | −0.03 | 0.481 | 0.001 |
| | BMI (25,30] | Women | −5.00 | 0.06 | 0.116 | 0.005 |
| | | Men | 1.48 | −0.06 | 0.143 | 0.004 |
| | BMI (30,35] | Women | −2.79 | 0.01 | 0.884 | 0.000 |
| | | Men | 6.10 | −0.13 | 0.069 | 0.017 |
| | BMI (35,Inf] | Women | −3.81 | 0.01 | 0.924 | 0.000 |
| | | Men | 21.07 | −0.41 | 0.130 | 0.066 |

**Table S12.** Regression coefficients and model statistics for the linear models (rate of change = intercept + slope ∗ age) in Figures S4–S23. The p-value is for the hypothesis test $H_0$ : slope = 0 vs $H_1$ : slope $\neq$ 0. BMI: body mass index, IDP: image-derived phenotype.
